# The Blood Proteome of Imminent Lung Cancer Diagnosis

**DOI:** 10.1101/2022.07.31.22277301

**Authors:** The Lung Cancer Cohort Consortium (LC3), Demetrius Albanes, Karine Alcala, Nicolas Alcala, Christopher I. Amos, Alan A. Arslan, Julie K. Bassett, Paul Brennan, Qiuyin Cai, Chu Chen, Xiaoshuang Feng, Neal Freedman, Florence Guida, Rayjean J. Hung, Kristian Hveem, Mikael Johansson, Mattias Johansson, Woon-Puay Koh, Arnulf Langhammer, Roger L. Milne, David Muller, Justina Onwuka, Elin Pettersen Sørgjerd, Hilary A. Robbins, Howard D. Sesso, Gianluca Severi, Xiao-Ou Shu, Sabina Sieri, Karl Smith-Byrne, Victoria Stevens, Lesley Tinker, Anne Tjønneland, Kala Visvanathan, Ying Wang, Renwei Wang, Stephanie Weinstein, Jian-Min Yuan, Hana Zahed, Xuehong Zhang, Wei Zheng

## Abstract

Identification of novel risk biomarkers may enhance early detection of smoking-related lung cancer. We measured 1,162 proteins in blood samples drawn at most three years before diagnosis in 731 smoking-matched case-control sets nested within six prospective cohorts from the US, Europe, Singapore, and Australia.

We identified 36 proteins with replicable associations with risk of imminent lung cancer diagnosis (all *p<4×10*^*-5*^). These included several documented tumor markers (e.g. CA-125/MUC-16 and CEACAM5/CEA) but most had not been previously reported. The 36 proteins included several growth factors (e.g. HGF, IGFBP-1, IGFP-2), tumor necrosis factor-receptors (e.g. TNFRSF6B, TNFRSF13B), and chemokines and cytokines (e.g. CXL17, GDF-15, SCF). The odds ratio per standard deviation ranged from 1.31 for IGFBP-1 (95% CI: 1.17-1.47) to 2.43 for CEACAM5 (95% CI: 2.04-2.89). We mapped the 36 proteins to the hallmarks of cancer and found that proliferative signaling, tumor-promoting inflammation, and activation of invasion and metastasis were most frequently implicated.

**Statement of significance:** After screening 1,162 proteins, we identified 36 markers of imminent smoking-related lung cancer diagnosis with a wide range of functions and relevance across the hallmarks of cancer. Forthcoming studies will address the extent to which these markers can discriminate future lung cancer cases and their utility for early detection.

## Introduction

Lung cancer is the leading cause of cancer death globally.^1^ The 5-year survival is 20%, but varies from 60% for early-stage disease (Stage I-II) to 6% for late-stage disease (stage IV).^2^ In the United States (US), lung cancer mortality declined by 6% annually from 2013 to 2016.^3^ This improvement can be attributed to advancements in diagnosis and treatment for patients with both early- and late-stage lung cancer.^4^ Improved surgical techniques, including stereotactic body radiotherapy (SBRT) and adjuvant chemotherapy, have improved prognosis for early-stage patients, whereas patients with locally advanced disease have benefitted from the introduction of radio-chemotherapy and adjuvant immunotherapy. However, most lung cancer patients are still diagnosed with late-stage disease where curative treatment is rarely possible, even though developments in targeted and immunotherapy combinations have improved short-term survival.^4^

Despite advances in lung cancer treatment, improving early detection is the most promising strategy to increase long-term survival. Screening with low-dose computed tomography (LDCT) has the potential to substantially increase the proportion of lung cancer patients diagnosed with early-stage disease who can be offered treatment with curative intent. The ability of LDCT screening to decrease lung cancer mortality among high-risk people with a history of smoking has been demonstrated in several randomized trials,^5,6^ but several concerns remain, including how to best identify and reach those individuals who are likely to benefit from screening, and how to manage indeterminate pulmonary nodules detected on LDCT.

The advent of LDCT screening and the introduction of targeted therapies have highlighted a need to identify lung cancer biomarkers that can be used to *i)* identify high-risk individuals who may benefit from screening, *ii)* inform diagnostic work-up and nodule management after LDCT screening, and *iii)* choose optimal treatment regimens and monitor response to treatment. In 2018, the US National Cancer Institute funded the Integrative Analysis of Lung Cancer Etiology and Risk (INTEGRAL) program, an ambitious initiative focusing on developing biomarkers that can refine eligibility criteria for LDCT screening and diagnostic work-up following LDCT.^7^ Here, we present results from the initial large-scale analysis designed to identify novel circulating protein biomarkers associated with imminent lung cancer diagnosis in the general population of smokers. Using a high-throughput proteomics approach, we screened over 1,000 circulating proteins in blood samples drawn up to three years prior to diagnosis within the Lung Cancer Cohort Consortium (LC3). We describe the proteins associated with lung cancer risk, the biological pathways to which they belong, and their known relevance in carcinogenesis.

## Methods

### Overview

Our study was designed to identify protein markers of imminent lung cancer in people with a smoking history from the general population. We defined imminent lung cancer as clinical lung cancer diagnosis within three years. Therefore, within six prospective cohort studies from the LC3, we identified 731 lung cancer cases who were diagnosed at most three years after blood draw, along with one smoking-matched control per case. We measured 1,162 proteins using the discovery platform from Olink Proteomics^8^ and assessed their association with lung cancer risk. We used a resampling-based validation algorithm to identify markers associated with lung cancer. We subsequently described this group of markers in more detail with respect to their epidemiological and gene expression characteristics, as well as their known relevance in carcinogenesis.

An overview of the analysis is presented in **Supplementary Figure 1**.

### Study sample

A detailed justification for the study design and description of the study sample is available in Robbins et al.^7^ In brief, we included six prospective cohorts of diverse geographical origin amongst cohorts participating in LC3. These included the European Prospective Investigation into Cancer and Nutrition (EPIC)^9^ from several countries in Europe, The Northern Swedish Health and Disease Study (NSHDS)^10^ from Sweden, the Trøndelag Health Study (HUNT)^11^ from Norway, the American Cancer Society Cancer Prevention Study-II (CPS-II)^12^ from the US, the Melbourne Collaborative Cohort (MCCS)^13^ from Australia, and the Singapore Chinese Health Study (SCHS)^14^ from Singapore. Lung cancer cases were eligible if they reported a current or former history of daily cigarette smoking at recruitment and were diagnosed with a histologically confirmed lung cancer (C34) at most three years after blood draw. Controls were selected by incidence density sampling and matched 1:1 to cases based on age (±1 year, relaxed to ±3 years for sets without available controls), date of recruitment (±1 month, relaxed to ±3 months), sex, and cohort, as well as smoking status in four categories (people who formerly smoked and quit <10 or ≥10 years prior, and people who currently smoked <15 or ≥15 cigarettes per day). The final study sample included 731 lung cancer cases and 731 matched controls. All research participants provided written, informed consent, and the study was approved by the relevant Institutional Review Boards.

### Proteomic measurements

Circulating proteins were measured using the discovery platform from Olink Proteomics.^15^ The Olink discovery platform is based on proximity extension assays (PEA) that are highly sensitive, avoid cross-reactivity, and have high reproducibility.^16^ Relative concentrations of up to 1,162 unique proteins, distributed over 13 Olink target panels, were measured by quantitative PCR (qPCR) (**Supplementary Figure 2**). Measurements are expressed as normalized protein expression (NPX) values which are log-base-2 transformed. Details on quality control metrics and coefficients of variation are available in the supplementary methods and **Supplementary Tables 1A and 1B**. Due to the high cost of Olink assays, we initially measured all 1,160 unique proteins only among the EPIC and NSHDS samples (n=252 case-control pairs), and then assayed the HUNT, CPS-II, SCHS and MCCS samples (n=479 case-control pairs) for a subset of promising panels which included between 392 and 484 proteins (see Robbins et al.^7^ and **Supplementary Figure 2**). For proteins measured on multiple panels within a single cohort (n=69 proteins with more than one measurement), we used the measurement with the highest variance and lowest missingness (see **Supplementary Methods**). Protein measurements were standardized within each cohort.

### Statistical analyses

The first step of our analysis aimed to identify proteins associated with imminent lung cancer diagnosis. Instead of using a single split-sample design, which can be subject to substantial influence from random chance, we applied a resampling-based validation algorithm which simulates a split-sample design repeated many times with many different random splits of the data. Specifically, in each of 500 iterations, we split the data into discovery (70%) and replication (30%) sets. In each of the 500 discovery and replication sets, we applied conditional logistic regression to estimate the odds ratio of lung cancer per standard deviation increment in relative concentration of each protein [OR_std_]. We applied this algorithm twice: once for the subset of 484 proteins measured in all 6 cohorts, and separately for the 678 proteins measured only in EPIC and NSHDS. In both algorithms, we balanced by cohort when splitting the data into random discovery (70%) and replication (30%) sets. In the algorithm including all 6 cohorts, we also ‘forced’ EPIC and NSHDS into the discovery set in every iteration since those data were used to choose the panels tested in the remaining 4 cohorts (**Supplementary Methods, Supplementary Figure 1**). Additional details on how missing protein data were handled during the resampling algorithm are in the **Supplementary Methods**.

We considered proteins to show replicable associations with imminent lung cancer if, in at least 50% of iterations, the *p*-value was below *p*<0.05/effective-number-of-tests (ENT)^17^ in the discovery set and below 0.05 in the corresponding replication set. The ENT method accounts for multiple testing by applying a Bonferroni correction, but determines the number of independent tests as the number of principal components needed to explain 95% of the variance in protein abundance.^17^

As a sensitivity analysis, we assessed the difference between the results of our resampling approach and a standard, single split-sample design. Here, we included only EPIC and NSHDS in the discovery set, since these data were used to choose the panels measured in the other 4 cohorts, which were defined as the replication set. We identified proteins that had a false-discovery-rate (FDR)-adjusted p-value<0.05 in the discovery set and a p-value<0.05 in the replication set. We chose the less conservative FDR significance instead of ENT significance because the power in the discovery set was lower due to smaller sample size.

For the group of markers identified as associated with imminent lung cancer by the resampling algorithm, we carried out additional analyses using the full dataset. For each marker, we calculated odds ratios for lung cancer stratified by histological type, stage, smoking status, cohort, and lead-time (time between blood draw and diagnosis) and examined trends by lead time (see **Supplementary Methods**). These stratified analyses did not account for multiple comparisons. To describe the association between each marker and smoking intensity, duration, and years since quitting, we used linear regression models fit among controls with adjustment for cohort, age, sex, and smoking status.

We also examined relationships between the identified markers. Separately among cases and controls, for pairs of proteins, we calculated Pearson’s correlation coefficients between the residuals of protein measurements after removing variance due to age, sex, and smoking status (‘residualized proteins’). To consider the relationships among all proteins simultaneously, we implemented sparse graphical network models. These models use a graphical LASSO-based resampling method on the partial correlations between residualized proteins to estimate a sparse set of connections among a set of proteins (see **Supplementary Methods**).^18^ We also identified biological pathways in which sets of connected proteins are enriched using g:Profiler.^19^

We subsequently evaluated the association between each identified marker and overall survival among cancer cases, separately using circulating blood measurements and tumor gene expression. For blood measurements, we applied Cox proportional hazards regression based on the time from lung cancer diagnosis to death from any cause, with stratification of the baseline hazard by cohort and sex and adjustment for age at recruitment. Models also included an interaction between lead time and the protein measurement, so that the coefficient for the protein is interpretable as its effect at the time of lung cancer diagnosis. For tumor gene expression, we extracted lung tumor RNA-seq gene expression for 480 adenocarcinoma and 420 squamous cell lung cancer patients from The Cancer Genome Atlas (TCGA) (see **Supplementary Methods**).

We compared cell specific expression of the markers (mRNA expression) in tissue extracted from cancer free individuals with that of tumor tissue. Expression data were extracted from the Human Protein Atlas^20^ and the Pathology Atlas.^21^ Details of these analyses are in the **Supplementary Methods**.

All statistical analysis were performed using R v4.1.2.

Finally, we used information available on GeneCards, the Human Protein Atlas, and Uniprot^22–24^ to match each protein’s function(s) to one or more of the Hallmarks of Cancer described by Hanahan and Weinberg^25,26^ in order to understand their biological roles.

## Results

The study sample included 731 incident lung cancer cases and 731 matched controls. Most study participants were men (980 vs. 482 women) and the mean age at blood collection was 65 years (standard deviation 9 years). The mean time between pre-diagnostic blood collection and diagnosis was 1.6 years (range: 0 to 3 years, by design) (**Table 1**). Demographic characteristics stratified by cohort are presented in **Supplementary Table 2**.

**Table 1.** Characteristics of 731 lung cancer cases and 731 matched controls from the Lung Cancer Cohort Consortium included in analyses to identify protein biomarkers of imminent lung cancer diagnosis.

| Characteristic | Cases<br>N (%) or mean (SD) | Controls<br>N (%) or mean (SD) |
| --- | --- | --- |
| Total number of participants | 731 | 731 |
| Female | 241 (33%) | 241 (33%) |
| Age, years | 64.8 (9.1) | 64.7 (9.2) |
| Body mass index, kg/m<sup>2</sup> | 25.5 (4.2) | 26.2 (4.3) |
| Follow-up time, years* | 1.6 (0.9) | 11.9 (5.4) |
| Follow-up survival time, years** | 4.1 (4.1) | 13.0 (5.4) |
| Smoking status |  |  |
| Current | 397 (54%) | 400 (55%) |
| Former | 334 (46%) | 331 (45%) |
| Cigarettes smoked per day | 20.9 (13.3) | 16.3 (11.7) |
| Years smoked | 39.5 (12.2) | 36.2 (14.0) |
| Years since cessation, among former | 15.4 (11.7) | 19.0 (13.6) |
| Histology |  |  |
| Adenocarcinoma | 246 (34%) |  |
| Squamous cell carcinoma | 150 (21%) |  |
| Large cell carcinoma | 27 (4%) |  |
| Small cell carcinoma | 118 (16%) |  |
| Other/NOS | 190 (26%) |  |
| Stage |  |  |
| Early stage (TNM 1/2) | 78 (23%) |  |
| Late stage (TNM 3/4) | 256 (77%) |  |
| Unknown/Missing | 397 |  |
| Participating cohort |  |  |
| CPS | 115 (16%) | 115 (16%) |
| EPIC | 188 (26%) | 188 (26%) |
| HUNT | 164 (22%) | 164 (22%) |
| MCCS | 108 (15%) | 108 (15%) |
| NSHDS | 64 (9%) | 64 (9%) |
| SCHS | 92 (13%) | 92 (13%) |

\*Time from blood draw to end of follow-up or lung cancer diagnosis
\*\*Time between specimen collection and the end of follow-up for mortality (including death)
Age, body mass index, and smoking information is assessed at the time of blood draw. EPIC: The European Prospective Investigation into Cancer and Nutrition; NSHDS: Northern Sweden Health and Disease Study; HUNT: The Trøndelag Health Study; MCCS: The Melbourne Collaborative Cohort Study; SCHS: The Singapore Chinese Health Study; CPS-II: The Cancer Prevention Study II

### Identification and description of proteins associated with imminent lung cancer

The associations between all 1,162 proteins and risk of imminent lung cancer diagnosis are reported in **Supplementary Table 3**. In the full study sample, there were 67 proteins associated with lung cancer after accounting for multiple comparisons (*p*<0.05/effective-number-of-tests) (**Supplementary Table 3**). Based on the resampling algorithm, 36 proteins were defined as associated with imminent lung cancer diagnosis (**Figure 1, Supplementary Figure 3, Supplementary Table 4**).

**Figure 1.**
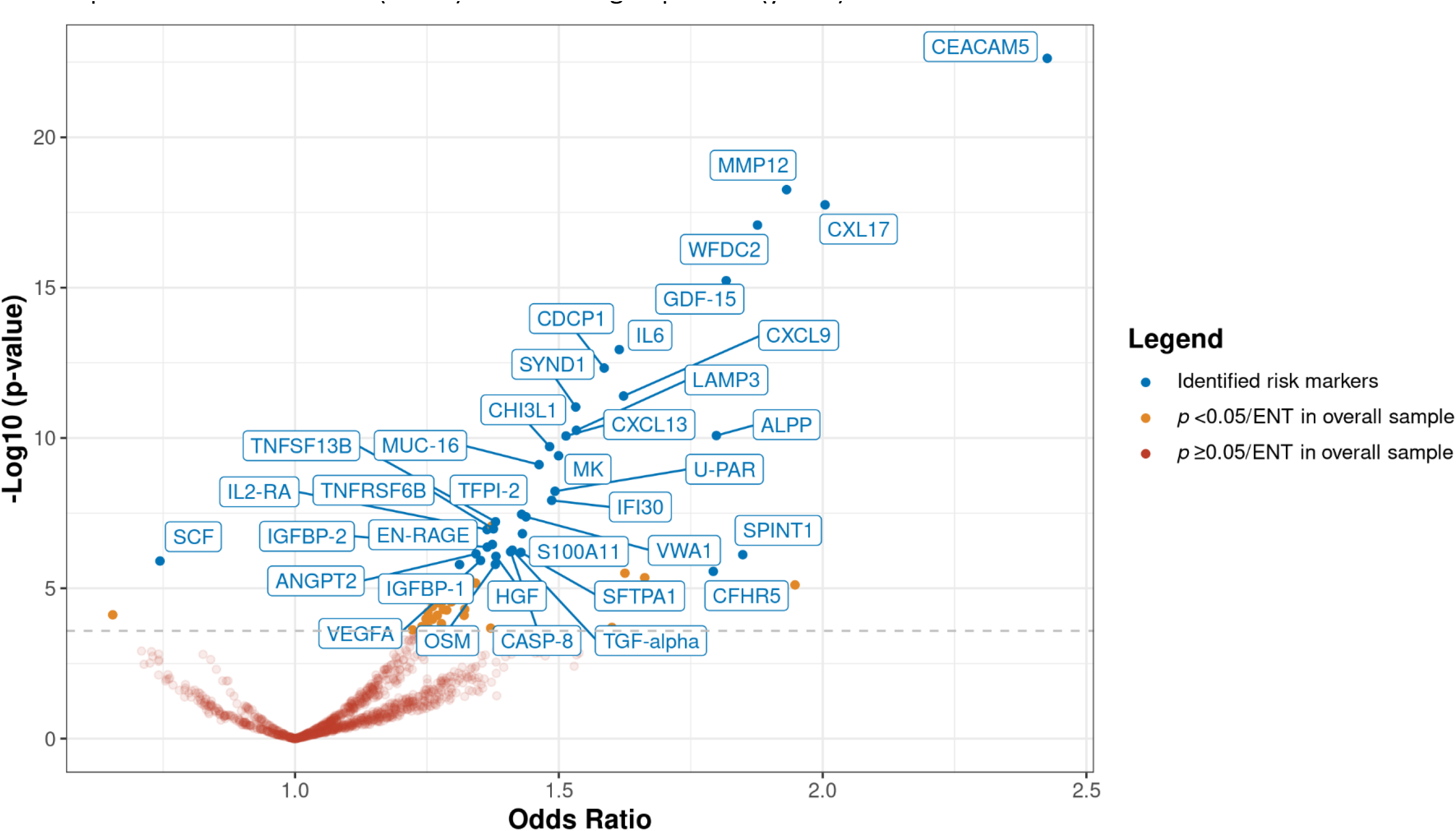
Identification of 36 protein biomarkers associated with risk of imminent lung cancer diagnosis among 731 cases and 731 matched controls in the Lung Cancer Cohort Consortium. The volcano plot depicts the lung cancer odds ratio per standard deviation increment in relative protein concentrations (x-axis) and the -Log10 p-value (y-axis). *The 36 markers of imminent lung cancer are labelled (see Methods). Markers were identified through a resampling process that measured the association of each protein with lung cancer risk in a discovery set and a replication set. The risk markers were required to have a p<0.05/effective-number-of-tests in the discovery set and p<0.05 in the replication set in at least 50% of the resampling iterations*.

**Supplementary Figure 4** compares the results of our resampling-based algorithm *versus* a single split-sample design to identify proteins associated with lung cancer. 29 markers were identified by both methods, 7 markers only by the resampling algorithm, and 10 markers only by the single split-sample method. Markers identified only by the resampling algorithm typically had larger ORs and smaller p-values in the full dataset compared with the proteins identified only by the single split-sample method. **Supplementary Table 5** shows that the proteins overlooked by the single split-sample method (e.g. CASP-8, S100A11) had weak p-values in the small discovery sets (respectively 0.06, 0.02) but strong p-values in the replication (2.1×10^−6^ and 4.6×10^−5^,respectively).

Among the 36 markers identified by the resampling algorithm, all but one showed a positive association with lung cancer (**Figure 1**). Among these, the estimated OR_std_ ranged from 1.31 (IGFBP-1, 95% confidence interval [95% CI]: 1.17-1.47, *p*=2×10^−6^) to 2.43 (CEACAM5, 95% CI: 2.04-2.89, *p*=2×10^−23^) **(Supplementary Table 3)**. The SCF protein was negatively associated with lung cancer (OR=0.74, 95% CI: 0.66-0.84, *p*=1.24×10^−6^).

Most of these markers displayed consistent associations with lung cancer across the major histological subtypes (**Supplementary Table 6**). Exceptions (*p*-heterogeneity [*p*_het_]<0.05) included CEACAM5, which was more strongly associated with adenocarcinoma than squamous cell carcinoma, and MMP12, which was more strongly associated with squamous cell carcinoma than with adenocarcinoma (**Supplementary Table 6, Supplementary Figure 5**). Two proteins (CXL17 and CEACAM5) showed stronger (*p*_het_<0.05) associations with risk of late-stage (T-stage 3-4) than early-stage lung cancer (T-stage 1-2) (**Supplementary Table 6, Supplementary Figure 6**). Stratification by smoking status highlighted two proteins, IGFBP-1 and VWA1, that had stronger associations among current than former smokers (*p*_het_<0.05, **Supplementary Table 6, Supplementary Figure 7**).

When stratifying by lead-time, 19 proteins showed stronger associations closer to diagnosis (*p*_het_<0.05, **Supplementary Table 6**) and 11 had a clear trend in the strength of association across categories of time since blood draw (*p*_trend_<0.05, **Supplementary Figure 8, Supplementary Table 7**). For instance, EN-RAGE displayed little evidence for an association with lung cancer more than one year prior to diagnosis (OR_2-3y_: 1.10, 95% CI: 0.91-1.33), but was strongly associated risk within one year of blood draw (OR_<1y_: 2.49, 95% CI: 1.87-3.32, *p*_het_=6×10^−6^). A similar pattern was observed for IL6 (OR_2-3y_: 1.36, 95% CI: 1.10-1.67 vs OR_<1y_: 2.56, 95% CI: 1.92-3.41, *p*_het_<0.001).

Analyses stratified by other demographic factors did not identify important heterogeneity in associations (**Supplementary Table 6**). However, in a separate exploratory analysis of study participants from the SCHS cohort (100% of participants are of Han-Chinese descent), we found two additional proteins (RFNG and S100A4, **Supplementary Figure 9**) associated with risk of lung cancer (*p*<0.05/effective-number-of-tests), despite showing little evidence among participants of European, US, or Australian cohorts. The OR_std_ for RFNG in SCHS was 2.65 (95% CI: 1.62-4.33) and 1.07 (95% CI: 0.93-1.23) in the other cohorts (*p*_het_<0.001), and the OR_std_ for S100A4 in SCHS was 2.77 (95% CI: 1.72-4.44) and 1.03 (95% CI: 0.90-1.18) in the other cohorts (*p*_het_<0.001).

When evaluating relationships with smoking history in a cross-sectional analysis among controls, we found that most markers differed markedly between former and current smokers, but only GDF-15 was associated with smoking intensity (**Supplementary Figure 10**). SCF was inversely associated with smoking duration and CXCL17 was positively associated with smoking duration. Further adjusting the main analysis for smoking intensity, duration, and body mass index (BMI) did not materially influence the OR estimates (**Supplementary Figure 11**).

### Relationships between proteins associated with imminent lung cancer

Pearson correlation coefficients between pairs of the 36 markers are depicted in **Supplementary Figure 12**, separately in cases and controls. Most proteins were moderately and positively correlated, except for SCF which was inversely correlated with some proteins (as well as with lung cancer risk, see above). Patterns were similar in cases and controls.

In the network analysis, the specific connections identified by sparse LASSO analyses among the 36 markers are shown separately for cases and controls in **Figure 2**. U-PAR was identified as the most highly connected and central protein in both the case and control networks (8 connections among cases and 9 among controls, **Supplementary Table 8**). Although most protein connections were common to controls and cases, we found evidence for three distinct clusters of proteins with stable associations observed only among cases. One was centered around SYND1 [Cluster_1_: U-PAR, IL2-RA, SYND1, HGF, and EN-RAGE] one around VEGFA [Cluster_2_: VWA1, VEGFA and IFI30], and one around MK and CXCL9 [Cluster_3_: MMP12, CXCL9, MK, and WFDC2]. The Cluster_1_ network was enriched for markers related to interleukin signaling and immune response (g:profiler pathway analyses *P*_adjusted_= 3×10^−2^ and 3×10^−3^, respectively) and Cluster_3_ was enriched for proteins involved in homeobox six-3 transcription factor (g:Profiler *P*_adjusted_: 4×10^−2^). Notably, several of the proteins most strongly associated with lung cancer, including CEACAM5, IL6, and SCF, were weakly correlated with other markers and did not have any stable connections (**Figure 2**).

**Figure 2.**
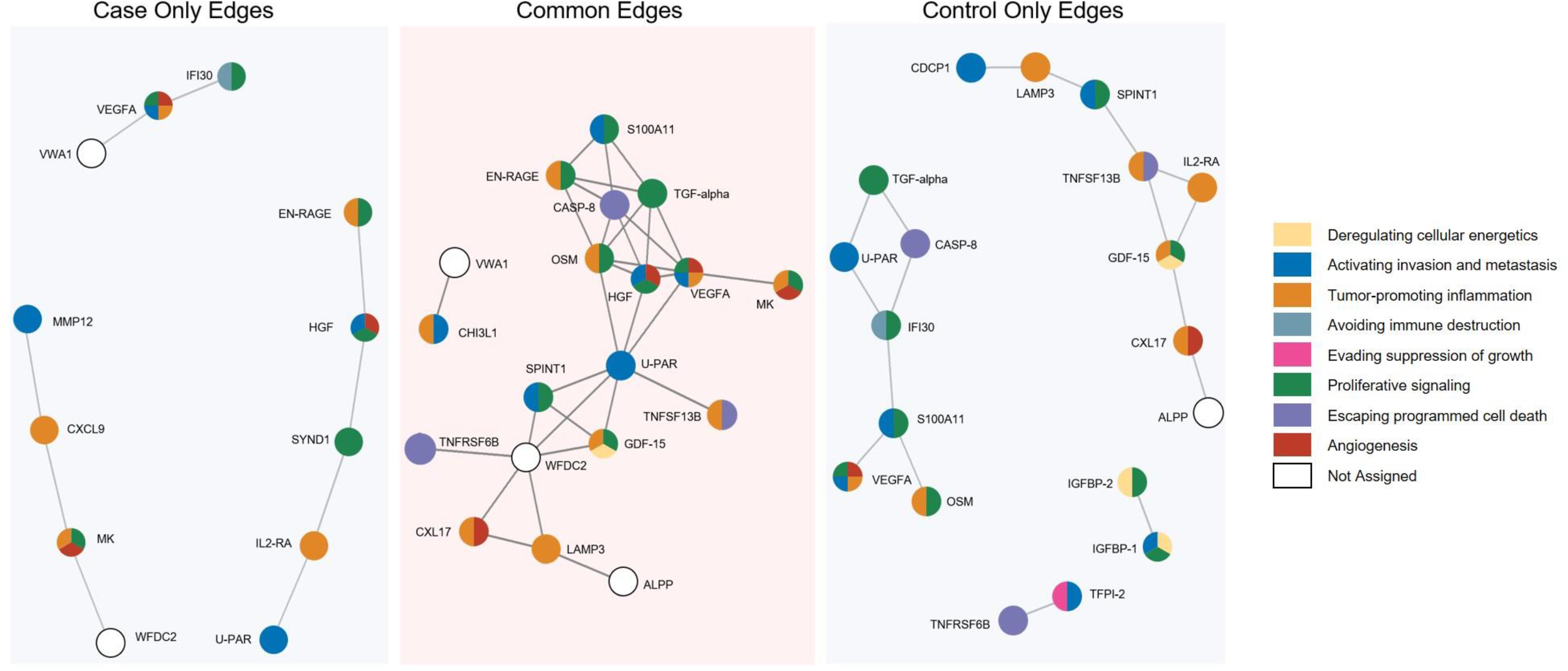
Network analyses among 36 protein biomarkers associated with risk of imminent lung cancer diagnosis. The figure depicts partial correlation networks and stable protein associations. *The partial correlation networks accounted for sex, age, cohort, and all other identified proteins. In lung cancer cases, no stable connections were found for ANGPT2, CDCP1, CEACAM5, CFHR5, CXCL13, IGFBP-1, IGFBP-2, IL6, MUC-16, SCF, SFTPA1, TFPI-2. In controls, no stable connections were found for ANGPT2, CEACAM5, CFHR5, CXCL13, CXCL9, IL6, MMP12, MUC-16, SCF, SFTPA1 and SYND1*.

### Associations with mortality among individuals with lung cancer

We evaluated the extent to which the 36 markers were associated with all-cause mortality following lung cancer diagnosis using both blood concentrations and tumor gene expression in TCGA samples. Whilst 20 proteins were nominally associated (*p*<0.05) with all-cause mortality when measured in blood (**Supplementary Figure 13**), these associations were relatively weak in comparison to the association with incident lung cancer risk. Of these 20 proteins, three were also associated with all-cause mortality when assessed using tumor gene expression (CDCP1, CEACAM5, and U-PAR) in TCGA.

### Gene expression in normal and tumor tissue

We used data from GTEx to assess mRNA expression for the 36 markers in normal tissue. Relative levels of mRNA expression in various normal cell types for 35 markers are shown in **Figure 3A** (data was not available for TNFRSF6B). Three markers (ALPP, SFTPA1, and MUC-16) were expressed primarily by lung cell types, while 4 others (IL2-RA, CXCL13, TNFSF13B, and EN-RAGE) were expressed primarily in immune cells. For mRNA expression in tumor cell types from TCGA, we found that most of the 36 markers were expressed in lung tumor tissue to some degree, but also in a wide variety of other cancer types (**Figure 3B**). The only marker that appeared specifically expressed in lung cancer tissue was SFTPA1.

**Figure 3.**
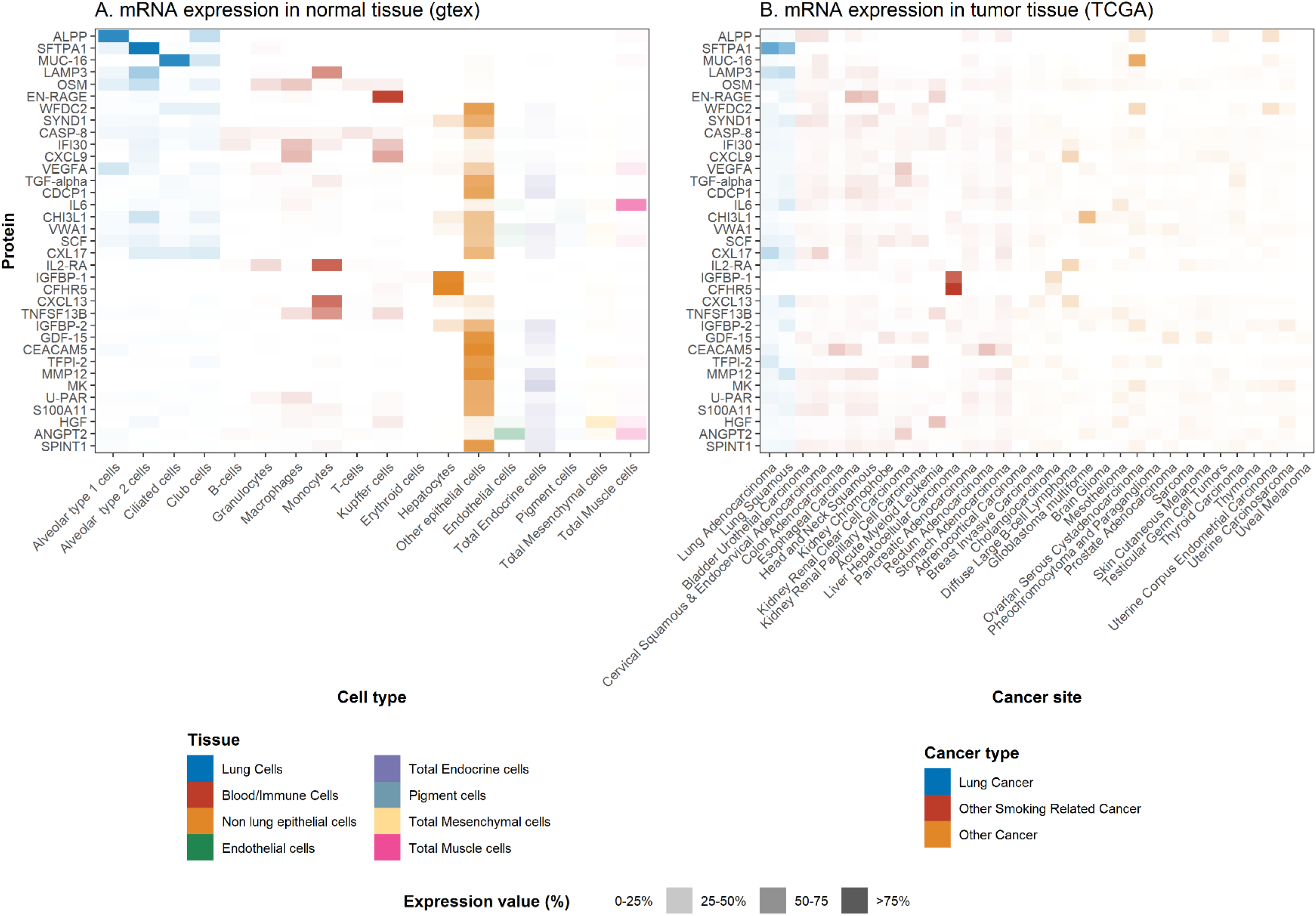
Gene expression of 36 protein biomarkers associated with risk of imminent lung cancer diagnosis in different types of normal tissue (A) and tumor tissue (B). *Proteins are listed in order of their relative expression in non-cancerous lung cells*.

## Discussion

The INTEGRAL project is a major initiative aiming to identify circulating protein biomarkers of imminent – but yet-to-be diagnosed – lung cancer. Based on blood samples drawn up to 3 years prior to clinical lung cancer diagnosis, we used a high-throughput proteomics platform to evaluate the association of 1,162 circulating proteins with imminent lung cancer in 731 cases and 731 matched controls from six prospective population cohorts. We identified 36 proteins associated with lung cancer diagnosis, most of which have not been previously identified as pre-diagnostic lung cancer biomarkers.

The last decade has seen major investments in research aiming to identify early cancer biomarkers. With the advent of early detection by LDCT screening, a strong focus has been placed on lung cancer. A wide array of circulating biomarkers have been proposed, including germline gene variants,^27,28^ microRNA,^29,30^ epigenetic markers,^31^ autoantibodies,^32^ protein markers,^33,34^ and circulating tumor DNA.^35^ However, few have been independently validated, and none are widely used in screening. In the INTEGRAL project, we decided to focus on circulating proteins due to their demonstrated ability to improve the discrimination of smoking-based risk prediction in an independent validation population,^34^ as well as the prospect of developing a clinical biomarker test at a reasonable cost and sample volume requirement.

Our current study analyzed 1,162 circulating proteins and found 67 proteins associated with imminent lung cancer after accounting for multiple testing. Following a resampling algorithm to simulate many iterations of split-sample discovery and replication, we identified 36 proteins associated with imminent lung cancer, 35 of which showed positive associations. At least eight of the 36 markers have been previously reported to be associated with lung cancer in pre-diagnostic samples, including several well-known tumor markers such as CEACAM5/CEA and CA-125/MUC-16,^34^ as well as IL6, CDCP1, CXCL9 and CXCL13.^36–38^ However, most of the 36 proteins are newly identified pre-diagnostic markers of lung cancer.

### Relevance of the identified proteins in cancer biology

The 36 identified markers have a wide range of molecular functions and include multiple growth factors (HGF, MK, IGFBP-1, IGFBP-2, MK, TGF-alpha, VEGFA), tumor necrosis factor-receptors (TNFRSF6B, TNFRSF13B), and chemokines and cytokines (CXL17, GDF-15, OSM, SCF). SCF, the only protein that we found to be negatively associated with lung cancer, has roles in regulation of cell survival and proliferation and hematopoiesis.^39^ The marker most strongly associated with lung cancer in our study was CEACAM5 (CEA), with a stronger association for adenocarcinoma than for squamous cell carcinoma. CEACAM5 is a surface glycoprotein that plays a role in cell adhesion, intracellular signaling and tumor progression^40^ and is routinely used to monitor recurrence among colorectal cancer patients.^41^ CEACAM5 was recently highlighted as a promising target for antibody-drug conjugate therapy of non-small cell lung cancer.^42^

To further contextualize the biological roles of the identified markers in cancer development, we assigned the proteins to one or more of the eight hallmarks of cancer as defined by Hanahan and Weinberg (**Figure 4**)^25,26^ based on their description and functions available on GeneCards, the Human Protein Atlas, and Uniprot.^22–24^ Amongst the 36 markers, 30 have documented functions within the hallmarks of cancer (**Figure 4**). We found that the most frequently implicated hallmark was proliferative signaling, which was associated with 15 markers, including growth factors such as HGF,^43^ TGF-alpha,^44^ and IGFBP-2,^43^ which are important in disease progression. Changes in proliferative signaling are common in lung tumors, as exemplified by the impact of deleterious mutations in well-described oncogenes, such as EGFR and KRAS.^45^ The second most frequently implicated hallmark was tumor-promoting inflammation, including markers such as CXCL9, CXCL13, CXL17, IL6, and IL2-RA. This highlights the central role for inflammation and the immune system in responding to the development of lung tumors. Third, unsurprisingly given frequent early development of metastases in lung cancer patients, many proteins mapped to activation of invasion and metastasis. Specifically, MMP12 and U-PAR have roles in the modulation of extracellular matrix (ECM) during metastasis.^46,47^

**Figure 4.**
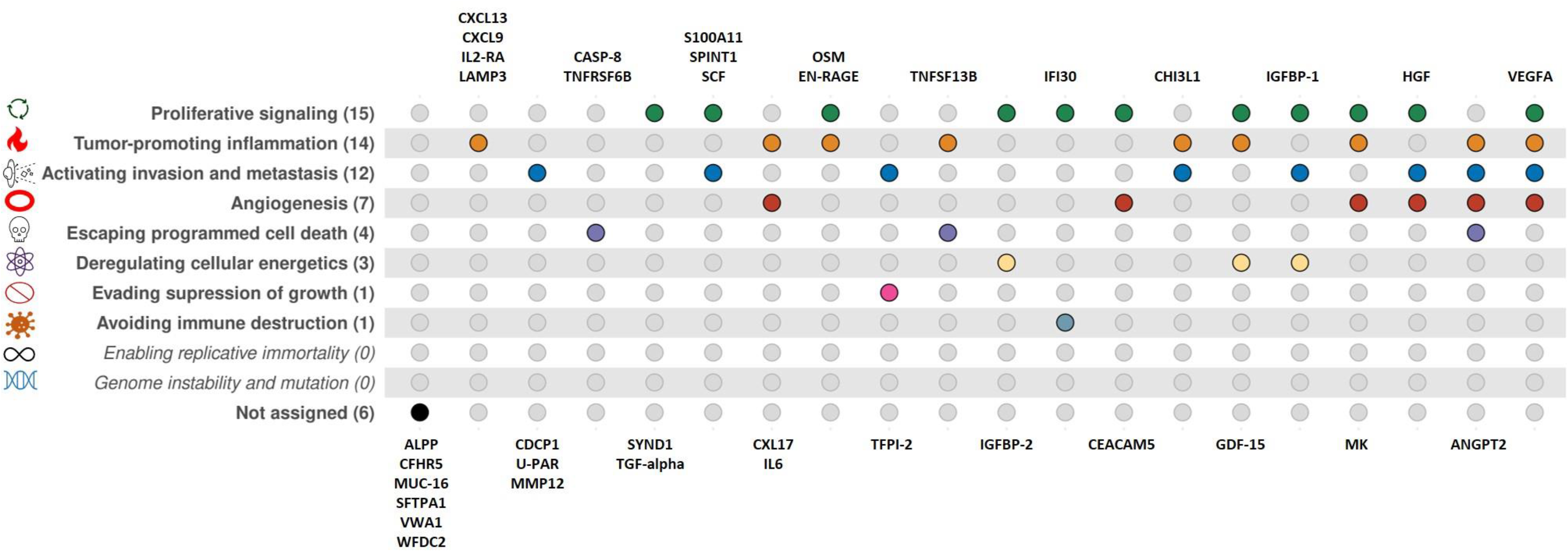
Relationship between 36 protein biomarkers associated with risk of imminent lung cancer diagnosis and the 10 hallmarks of cancer. *We used protein descriptions and functions available on GeneCards, the Human Protein Atlas, and Uniprot to assign the 36 markers to one or more of the eight hallmarks of cancer as defined by Hanahan and Weinberg*.

To understand why circulating concentrations of the identified proteins are associated with lung cancer onset, and to assess whether they are likely to be specific to lung cancer – as opposed to cancer at other sites – we used publicly available expression data for a range of normal and tumor tissues. This analysis yielded two notable observations; first, that only three proteins, ALPP, SFTPA1, and MUC-16, were predominantly expressed in normal lung cells compared to cell types of other origins. In contrast, several proteins appeared to be primarily expressed by immune cells, although most were also expressed by other cell types. The second notable observation was that only one protein – SFTPA1 – was predominantly expressed by lung tumor tissue compared to other tumor tissues, whereas most proteins were expressed in a wide range of cancer types. These complementary data suggest that few of the identified markers are likely to have originated in yet-to-be diagnosed lung tumor tissue, but rather are present in the circulation as a somatic response to subclinical cancer.

### Epidemiological properties of identified proteins

Many of the 36 markers displayed stronger associations when measured in blood drawn closer to diagnosis. This is expected for markers indicative of forthcoming disease, as opposed to markers of disease etiology. Among these proteins, two from the S100 family (EN-RAGE and S100A11) displayed particularly strong associations closer to diagnosis. Proteins in the S100 family have previously been proposed as biomarkers for prognosis of malignant melanoma.^48,49^

We found limited evidence for heterogeneity in associations for most of the 36 markers when stratifying by smoking status, and little impact of additional adjustment for smoking characteristics. This suggests that the associations reflect a somatic response to (or the direct action of) a subclinical lung tumour, rather than differences in tobacco exposure. Encouragingly, this also implies that the markers have potential to improve short-term lung cancer risk prediction beyond existing risk calculators that leverage detailed information from smoking history.^33,34,50^

To better understand the relationships between the 36 markers, we conducted a sparse graphical LASSO-based network analysis and observed specific associations between 12 proteins among cases that did not appear among controls. These case-specific protein connections were clustered in three groups and were all broadly implicated in an extracellular defense response to somatic stress. In contrast, connections that were specific to controls appeared to be more strongly associated with a signaling response to cell proliferation. In seeking to establish a risk prediction model including multiple proteins, we would anticipate some redundancy in the risk discriminative performance of connected proteins.

Finally, we found that 20 of the 36 identified markers were also associated with increased risk of all-cause mortality following lung cancer diagnosis when measured in blood. A subset of these markers (U-PAR, CEACAM5, and CDCP1) were also associated with all-cause mortality with concordant direction in an independent sample when measured as mRNA in lung tumor tissue. This may suggest a role for these markers in tumor progression or an immune or inflammation response in lung tissue. Notably, CDCP1 was previously associated with an increased risk of lung cancer in pre-diagnostic blood,^38^ is overexpressed in lung cancer tissue,^51^ and is associated with metastases and poor prognosis.^51–54^

### Strengths and limitations

Our results are based on protein data generated for the specific purpose of identifying novel early detection markers of lung cancer. The study design, with pre-diagnostic samples drawn up to 3 years prior to clinical (non-screening detected) lung cancer diagnosis, ensured that identified markers were not influenced by the diagnosis itself or subsequent treatment, as in a retrospective case-control study of diagnosed cases.^55^ By drawing samples from multiple studies, we were able to verify the consistency of associations across populations from the US, Europe, Southeast Asia, and Australia. Furthermore, our sample size provided 80% power to identify markers with an OR_SD_ of at least 1.26 after considering multiple testing, suggesting it is unlikely that we failed to identify any marker among the 1,162 proteins that is of major use for early detection. Future discovery studies seeking to identify protein markers for early lung cancer detection may therefore consider using our results as an initial reference and focus additional investments on measuring non-overlapping sets of markers.

An important limitation of our study was that information on clinical stage was lacking for most cases (**Table 1**). This limited our ability to evaluate whether the identified markers were primarily driven by lung cancer diagnosed at late stage. However, based on the available stage information, we did not observe systematic differences between the OR estimates for early vs. late stage lung cancer (**Supplementary Table 6**).

Our controls were sampled directly from the same source population as cases and were individually matched to cases by detailed smoking characteristics, age, sex, and timing of the blood draw. This design protects against multiple types of bias that frequently affect biomarker studies. However, our nested case-control design does not readily allow us to establish absolute risk models, nor to evaluate the utility of our markers for risk prediction in the general population, because such metrics are strongly influenced by the highly selected controls. As described by Robbins et al.,^7^ we will address this question in a large, independent validation phase by analyzing pre-diagnostic blood samples from a larger sample of 1,700 lung cancer cases and 2,900 randomly selected cohort representatives including 10 additional cohorts participating in the Lung Cancer Cohort Consortium.

### Future directions

In future work, we plan to study the dynamics of the identified markers by evaluating repeat blood samples collected from the same individuals over time. As the majority of our study population was of European descent (with the exception of the majority Han Chinese participants in the SCHS cohort), an important future aim is to determine whether any additional markers might be important specifically for populations of non-European ancestry. In addition, our study focused explicitly on people with a smoking history, and we consider it unlikely that the most relevant set of markers for lung cancer among people who never smoked were identified. Finally, we note that there is substantial scope for future studies to explore the potential biological roles of the identified markers in lung cancer development and progression.

## Conclusion

After screening 1,162 proteins, we identified 36 markers of imminent lung cancer with a wide range of functions and relevance across the hallmarks of cancer. Forthcoming studies will address the extent to which these markers can discriminate future lung cancer cases and their utility for early detection. Our study provides a first expansive view of the blood proteome in the years leading up to diagnosis of smoking-related lung cancer and can serve as a reference for investigations seeking to identify early protein markers of lung cancer.

## Data Availability

All data produced in the present study are available upon reasonable request to the PIs.

## Disclaimer

Where authors are identified as personnel of the International Agency for Research on Cancer / World Health Organization, the authors alone are responsible for the views expressed in this article and they do not necessarily represent the decisions, policy or views of the International Agency for Research on Cancer / World Health Organization.

## Conflicts of interest

None

## Funding

This study was supported by the US NCI (INTEGRAL program U19 CA203654 and R03 CA245979), Fondation ARC pour la recherche sur le cancer and l’Institut National Du Cancer (INCa) (INCA201601246/ARC_10450, Fondation ARC et INCa, France), INCa (TABAC18-035, France), the Cancer Research Foundation of Northern Sweden (AMP19-962), an early detection of cancer development grant from Swedish Department of Health ministry, and Cancer Research UK [C18281/A29019]. RJH is supported by the Canada Research Chair of the Canadian Institute of Health Research.

## Author contributions

**In alphabetical order:**

**Join first authors/contributed equally:** KA, FG, MiJ, KS-B, VS and HZ

**Drafted manuscript:** MiJ, MaJ, HAR, KS-B, VS and HZ

**Statistical analyses:** KA, XF, FG, JO, KS-B, and HZ

**Read and approved manuscript:** All authors

**Supervision:** MaJ and HAR

**Provided administrative support:** DA, AAA, JKB, PB, QC, CC, NF, KH, MiJ, MaJ, W-PK, AL, RLM, EPS, HAR, HDS, GS, XOS, SS, VS, LT, AT, KV, YW, RW, SW, J-MY, XZ and WZ

## Acknowledgments

We would like to thank Matthieu Foll and Lynnette Fernandez Cuesta at the International Agency for Research on Cancer (IARC/WHO) for their valuable contribution to our understanding and interpretation of the results in this study.

## Notes

### Competing Interest Statement

The authors have declared no competing interest.

### Author Declarations

IARC's (International Agency for Research on Cancer) ethics committee

