## Supplementary Files for "The Blood Proteome of Imminent Lung Cancer Diagnosis"

### Supplementary Methods

#### Proteomics Measurements

Circulating proteins were measured using the Olink Proteomics multiplex platform (Uppsala, Sweden).^1^ The technology is based on a proximity extension assay (PEA) technique that is highly sensitive and avoids cross-reactivity with high reproducibility. The full protocol of the PEA has been reported previously.^2^ The proteins are allocated across 13 separate panels, each including 92 proteins focused on a specific area of disease or biology. All sample plates included four internal control samples to monitor the quality of assay performance, as well as the quality of individual samples. The quality control (QC) was performed in two steps:

1. The standard deviation of the internal controls was evaluated for each sample plate. Only data from sample plates with a standard deviation below 0.2 NPX were considered valid.

2. The quality of each sample was assessed by evaluating the deviation from the median value of the controls for each individual sample. Only samples that deviated less than 0.3 NPX from the median passed the quality control. Fewer than 5 samples failed this quality control check.

We initially used all panels to measure 1,160 unique proteins on samples from EPIC and NSHDS (n=252 case-control pairs, some proteins were measured on several panels and the total number assays was 1,290). As outlined by Robbins et al.,^3^ because of the incremental cost implications of applying each additional Olink panel, we selected five to six panels (392- and 484 proteins) based on the results from EPIC and NSHDS data that were assayed on the remaining samples from HUNT, MCCS, CPS-II and SCHS (n=478 case-control pairs). Details on the panels measured for each cohort are available in **Supplementary Figure 2**. The panels focus on proteins with relevance for different processes such as immunity (e.g. inflammation), cell regulation (e.g. regulation of cell proliferation, cell death/apoptosis), tissue generation and remodeling (e.g. angiogenesis, heart development), as well as mechanisms that are central to the initiation and progression of diseases such as cancer (e.g. angiogenesis, cell differentiation and adhesion) and neurology-related diseases. Pairs of case-control samples were plated together, with the pairs randomly allocated over 96-well plates. Protein concentrations were measured by quantitative PCR (qPCR) to quantify relative protein concentrations expressed as normalized protein expression (NPX) values on the log2 scale. Measurements below the lower limit of detection (LOD) were replaced with the LOD divided by the square-root of 2.^4^

Overall, 112 proteins were measured on more than one panel, with some proteins assayed on 5 different panels. For analysis, we chose one measurement per protein by cohort. First, for each protein we prioritized the measurements from the four panels measured on all cohorts (Cardiovascular III, Immuno-Oncology, Inflammation, and Oncology II). Then, if needed, we selected the measurement with the highest variance within each cohort. Data on selected assays by protein by cohort are described in **Supplementary Table 1**. Protein measurements were standardized by cohort.

#### Statistical Analyses

Our resampling algorithm is described in the main text. Here, we provide additional details about its implementation.

To account for missing protein data (specifically when dealing with a protein that is not measured in a cohort), at each iteration for each protein we removed individuals with missing values for the protein in question before splitting the data into discovery and replication. Protein measures were rescaled at each iteration by cohort, separately for the discovery and replication set.

Because many proteins were only measured in the EPIC and NSHDS cohorts, to identify the markers associated with lung cancer, we applied the resampling algorithm twice: once on EPIC and NSHDS alone (n=678 proteins), and once on all cohorts together (n=484 proteins) (see **Supplementary Figure 1**). Case-sets were allocated randomly into discovery (70%) and replication (30%) while balancing by cohort. We calculated the effective number of tests separately for the two sets of proteins.

To obtain preliminary estimates of the improvement in discrimination provided by each single protein beyond the PLCO*_m2012_* risk model, we first calculated each participant’s *PLCO_m2012_* risk score and estimated the AUCs. We note that these AUCs, calculated in data from matched cases and controls, are artificially reduced compared with AUCs that would be obtained in a representative population. Then, for each protein, we fit a conditional logistic regression model in the overall data with an offset for the natural logarithm of the *PLCO_m2012_* risk score. We predicted the *PLCO_m2012_+protein* risk score in the same dataset and estimated the AUC of that integrated model. We calculated the improvement in discrimination provided by each protein as the difference in AUCs from each model. We used the pROC package in R to calculate the AUCs. The AUC results are not discussed but can be found in **Supplementary Table 5**.

For the 36 proteins identified by the resampling algorithm, we assessed trends by pre-diagnostic lead time in the association between each protein and lung cancer risk. Here, we report the beta coefficient, p-value, and Z-score of the interaction between the protein and lead-time from a conditional logistic regression model additionally adjusted for the protein measurement.

#### Protein Correlations and Network Analyses

To evaluate how the identified proteins associate with each other distinctly among cases and controls, we first removed variation in protein concentrations due to age, sex, and smoking status by taking residuals from a linear regression of each protein (separately) on these three factors. We subsequently calculated Pearson’s correlation coefficients among proteins, separately among cases and controls, and presented correlations with *p*<0.05.

To consider relationships between all markers associated with lung cancer simultaneously, we employed sparse graphical network models that estimate the network topology of protein correlations. We estimated these networks based on data from EPIC and NSHDS only to include all identified proteins. As for the pairwise correlation analysis, we applied the sparse graphical network models on the residuals from a linear regression of each protein on age, sex, and smoking status. We estimated the sparse network for the identified markers separately in cases and controls.

The sparse graphical network models use a graphical LASSO-based re-sampling method on the partial correlations between proteins to estimate a sparse set of connections between a set of proteins.^6^ It has three main parameters: the LASSO penalization parameter (λ) to determine the degree of sparsity in the network, the threshold for the proportion of resamplings (π) where a given connection between two proteins is observed, and the per-family error rate (PFER) to determine a ceiling on the number of false protein-protein connections in a network. We set the PFER to < 5% of the total potential network size [N*×*(N-1)/2]. Once the PFER is set, we chose values for λ and π by maximizing the negative log-likelihood estimator of the stability of the network. The resulting networks in cases and controls can be interpreted as the sparse and stable set of adjusted protein-protein connections, without direction.

We subsequently used these networks to identify the protein-protein connections that were common in control and case networks, that were unique to controls, and that were unique to cases. We considered network statistics including normalized group-level centrality, as a measure of how structured the network of associations were around a central group of important proteins, calculated using the igraph package on R^7^.

#### All-Cause Mortality

##### Association of tumor gene expression with all-cause mortality among lung cancer cases

For tumor gene expression analyses of the identified markers, we extracted lung tumor RNA-seq gene expression for 480 adenocarcinoma and 420 squamous cell lung cancer patients from The Cancer Genome Atlas (TCGA) project 2731 via dbGAP. We applied Cox regression to estimate hazard ratios for all-cause mortality based on a standard deviation increment in gene-expression. These models included stratification of the baseline hazard by sex and histological subtype and adjustment for age at diagnosis.

#### Tissue and Tumor Expression Proteins

Single cell mRNA expression data available through the Human Protein Atlas^8^ was used to describe mRNA expression for genes that code for the identified markers taken from cancer-free individuals. Normalized expression levels were obtained using single cell RNA sequencing of 51 cell types from 13 different human tissues. Cell-specific expression was calculated as the ratio of each cell type expression to the total expression across all cell-types for each gene. We included epithelial, endocrine, endothelial, muscle, pigment, mesenchymal, and blood and immune cells. Expression in neuronal, glial, germ, trophoblast, and undifferentiated cells was not included because these tissues were unlikely to contribute significantly to circulating levels of these proteins in adult men and women. In **Figure 3**, individual cell type expression levels are shown for 4 lung-specific epithelial cell types, 7 blood and immune cell types, hepatocytes, endothelial cells, and pigment cells. Summed expression levels are shown for the 15 remaining epithelial cell types (labeled other epithelial cells), the 3 endocrine cell types, 3 mesenchymal cell types, and 2 muscle cell types. Expression was defined to be minimal for cell types with <5% of the total mRNA expressed.

The same methodology was applied to mRNA expression data from the Pathology Atlas^9^ to quantify expression by various tumor types.

### Supplementary Tables

*Supplementary Tables are found in the supplementary Excel file. Each is present on a different sheet with the table number. Sheets are ordered.*

Supplementary Table 1a and Supplementary Table 1b: Quality controls of assay measures in the EPIC and NSHDS cohorts (1a), and in in the CPS, HUNT, MCCS and SCHS cohorts (1b).

Supplementary Table 2: Characteristics of 731 lung cancer cases and 731 matched controls stratified by cohort.

Footnote: Follow-up time for lung cancer may be shorter than follow-up time for mortality due to different end dates for the completeness of cancer registry vs mortality registry data.

Supplementary Table 3: Observed effect size of all measured proteins with lung cancer risk in the full data.

Supplementary Table 4: Proportion of 500 random discovery-replication samples in which risk-associated proteins were replicated.

Supplementary Table 5: Comparison of the estimated associations between each protein and lung cancer risk identified by the single split design vs the resampling algorithm.

Supplementary Table 6: Stratified associations of the 36 identified markers with lung cancer risk across different strata.

Supplementary table 7: Trends by lead time in the association between the 36 identified markers and lung cancer risk.

Footnote: we defined lead time as the time (in years) elapsed between blood draw and clinical diagnosis of lung cancer.

Supplementary Table 8: Centralities of the penalized networks of the 36 identified markers.

Footnote: Degree centrality represents the number of edges each node has (i.e. the number of proteins each protein is directly connected to).

Betweenness centrality represents the importance of each node to the flow of the network by assessing the number of short paths between two nodes each node is on (i.e. if protein A is connecting protein B and C and there’s no other link to B and C, then A would have a high betweenness centrality. If protein D is connected to B and has no other connection, D is therefore not linking any proteins and will have a low betweenness centrality).

Closeness centrality represents the average distance between each node and the other nodes (i.e. for each protein we calculate the inverse of the sum of the distances to every other protein). The higher the closeness is, the more each protein is efficiently related to the other proteins in the network.

Eigen vector centrality is an extension of degree centrality. It adjusts the centrality degree assigned by the number of direct links to each protein for their “power”. In other words, if protein A and protein B are each independently linked to 5 other proteins then both have a high degree centrality. However if the 5 proteins linked to protein A have no importance in the network (are not linked to other proteins) while the 5 proteins linked to B are linked to other proteins in the network than protein B will have a higher eigenvector centrality than protein A.

### Supplementary Figures

Supplementary Figure 1**:** Flow chart summarizing our method to identify protein markers associated with imminent lung cancer diagnosis.

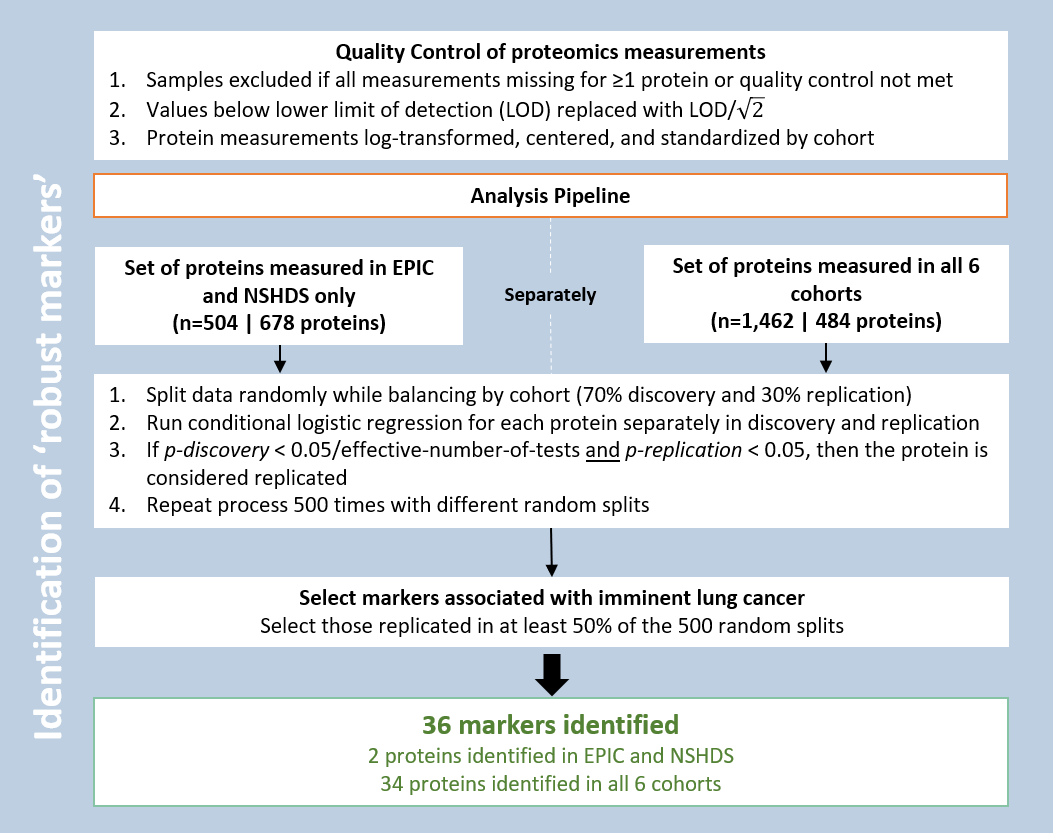

EPIC: The European Prospective Investigation into Cancer and Nutrition; NSHDS: Northern Sweden Health and Disease Study.

HUNT: The Trøndelag Health Study; MCCS: The Melbourne Collaborative Cohort Study; SCHS: The Singapore Chinese Health Study; CPS-II: The Cancer Prevention Study II.

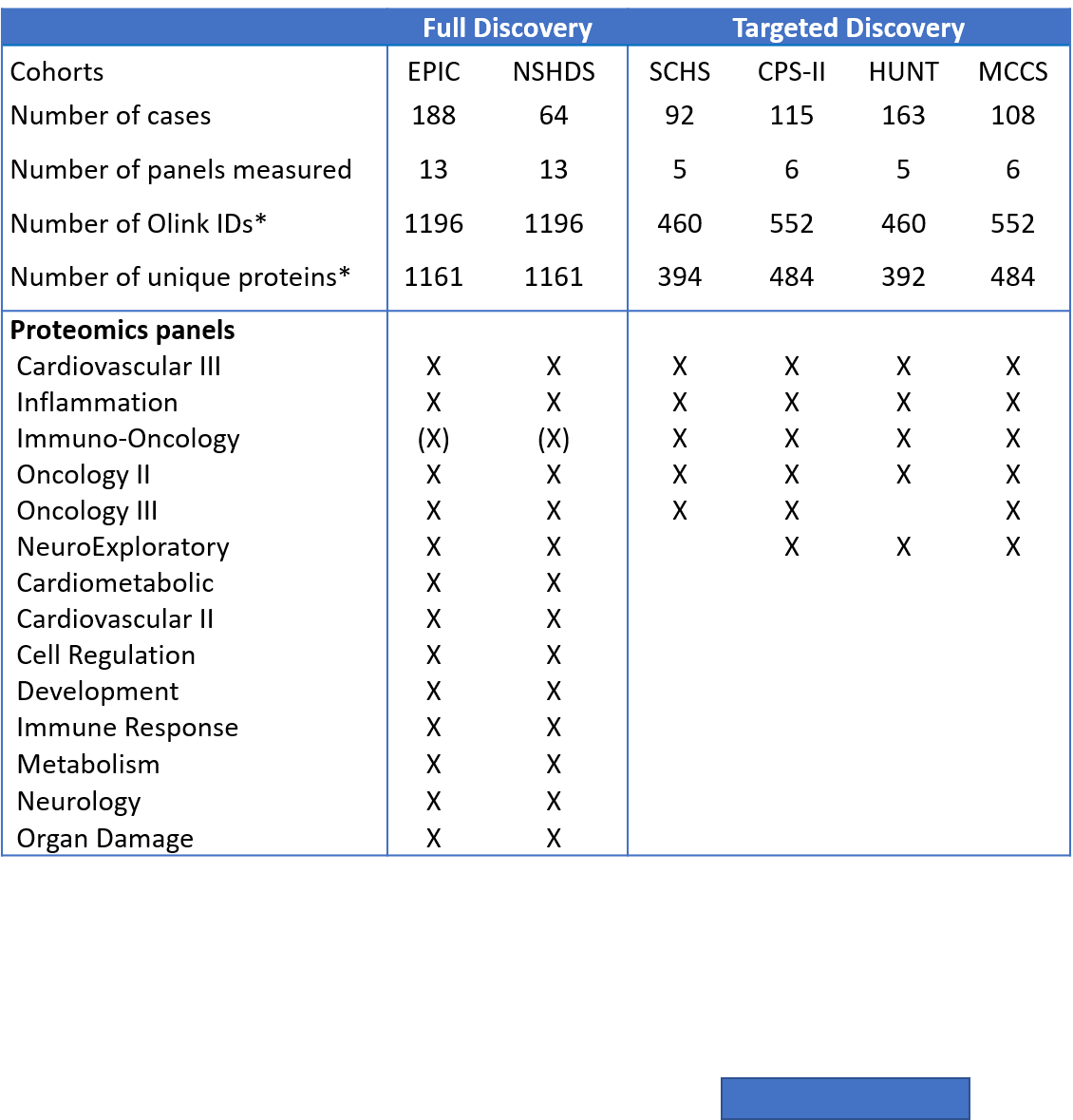
Supplementary Figure 2**:** Description of panels measured within cohorts (*Robbins et al, 2022,* https://doi.org/10.1101/2022.03.21.22272544)

*Some proteins are measured on multiple panels and therefore have multiple Olink IDs for the same protein. In these cases, for each protein, we chose a single Olink ID for analysis by choosing the one that was measured on more cohorts, and then if needed, the Olink ID with the highest variance.

(X): all the proteins from the Immuno-Oncology panel are included on other panels assayed as indicated.

Overall, 1163 unique proteins were measured by Olink. 1161 is the number of unique proteins measured in EPIC and NSHDS (2 proteins from the NeuroExploratory panel (ADGRB3 and LTBP3) were not measured in EPIC and NSHDS but had measurements in the remaining cohorts. Throughout the manuscript we refer to 1162 proteins analyzed, as 1 protein (MAPT) was excluded from the analysis due to invariant measurements (standard deviation = 0) in all cohorts, and refer to n=1160 proteins analyzed in EPIC and NSHDS.

EPIC: Investigation into Cancer and Nutrition, NSHDS: The Northern Swedish Health and Disease Study (NSHDS), HUNT: the Trøndelag Health Study, CPS-II: the American Cancer Society Cancer Prevention Study-II, MCCS: the Melbourne Collaborative Cohort, SHCS: Singapore Chinese Health Study (SCHS).

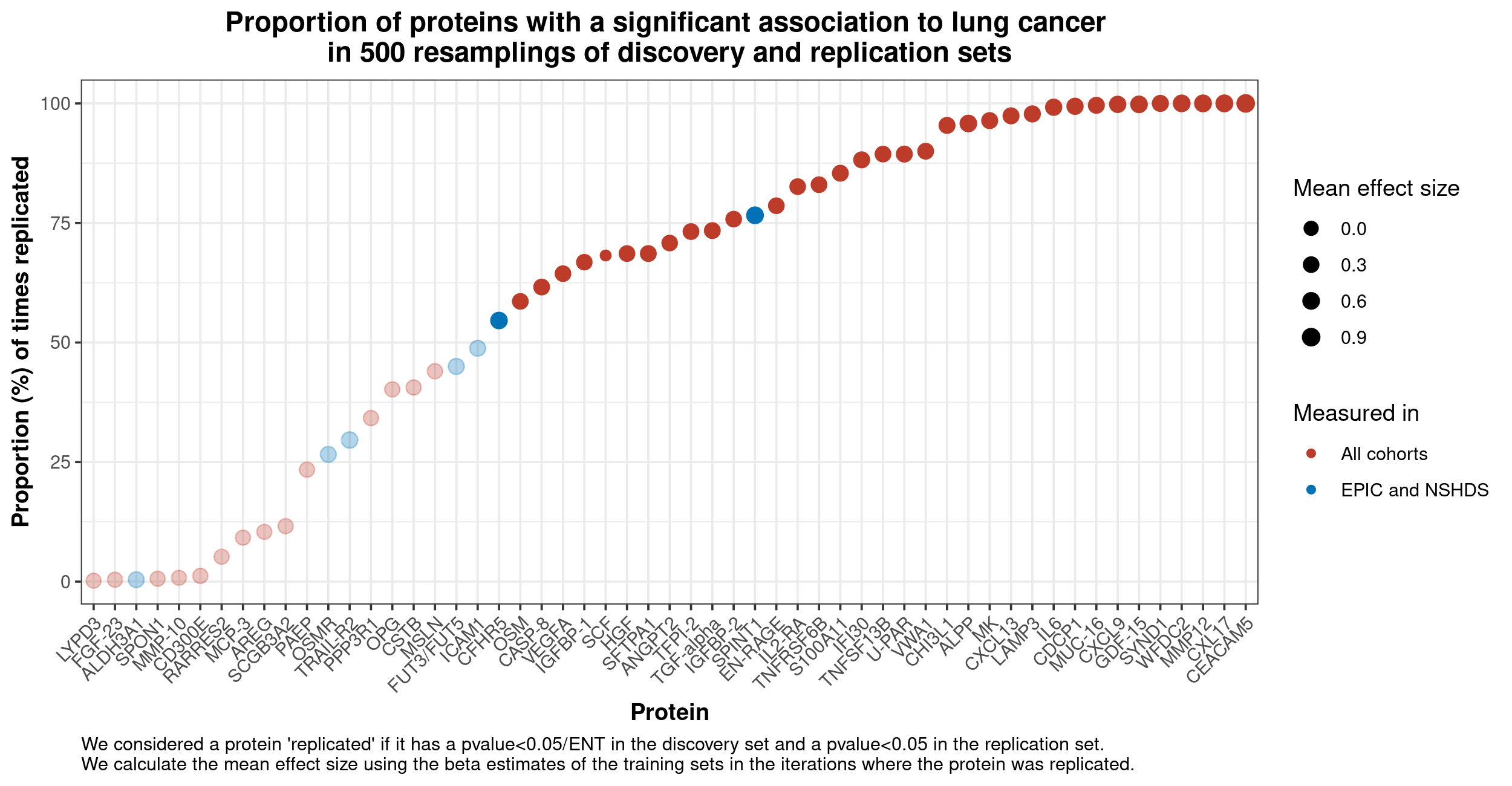
Supplementary Figure 3: Proportion of 500 random discovery-replication samples in which risk-associated proteins were replicated.

We defined replicated biomarkers as biomarkers with an association below p<0.05/ENT in the discovery set and p<0.05 in replication set in one iteration. We calculated the mean effect size using beta estimates in training sets of the iterations where the protein was replicated. Mean effect size can be found in **Supplementary Table 4**.

**Supplementary Figure 4**: Comparison of the estimated associations between each protein and lung cancer risk identified by the single split design vs the resampling algorithm.

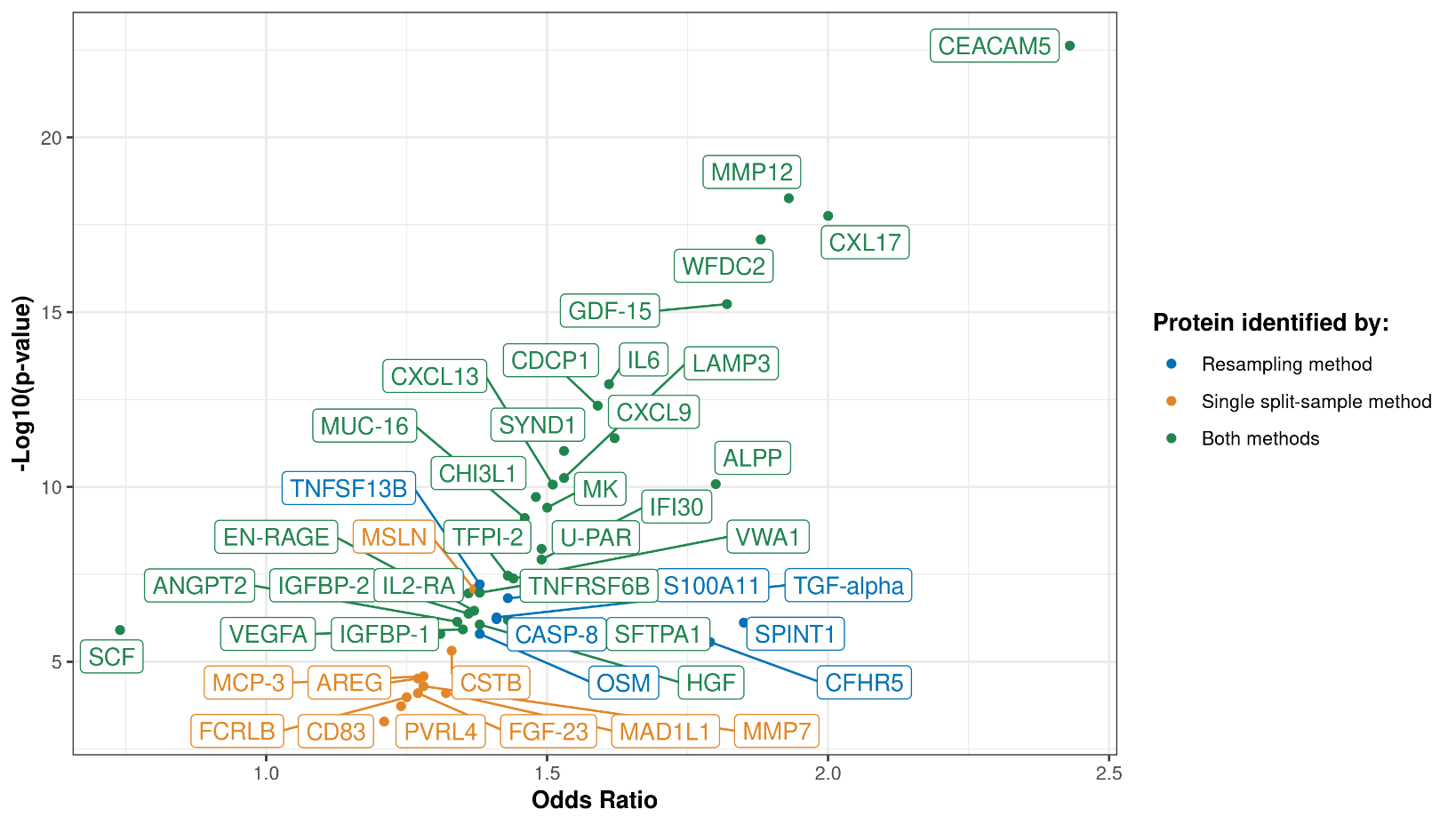

Supplementary Figure 5**:** Heterogeneity in risk-associations for MMP12 and CEACAM5 by lung cancer histological type (adenocarcinoma vs squamous cell carcinoma, *p*_het_ <0.05).

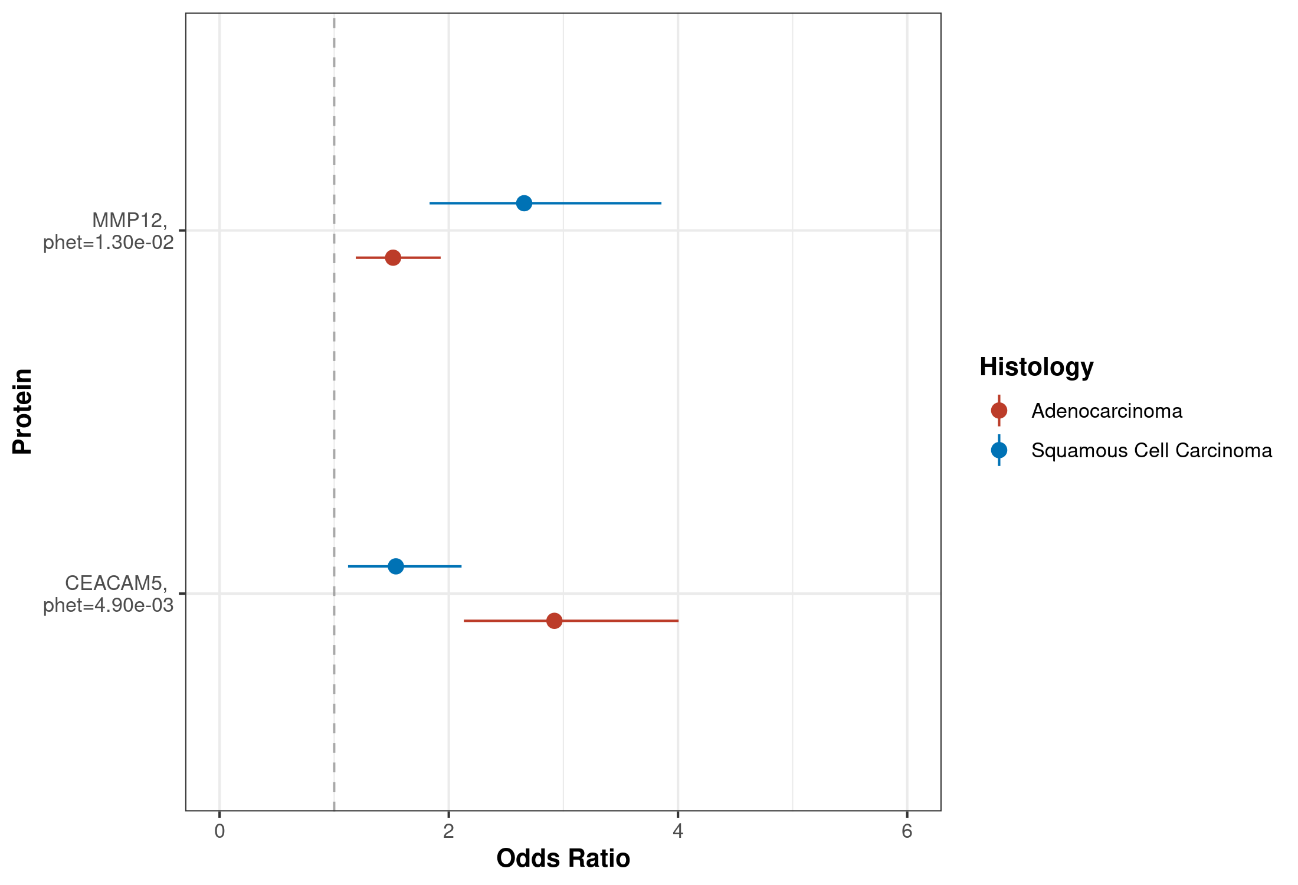

Supplementary Figure 6**:** Associations between the 36 identified and lung cancer risk, stratified by lung cancer stage (I/II vs III/IV).

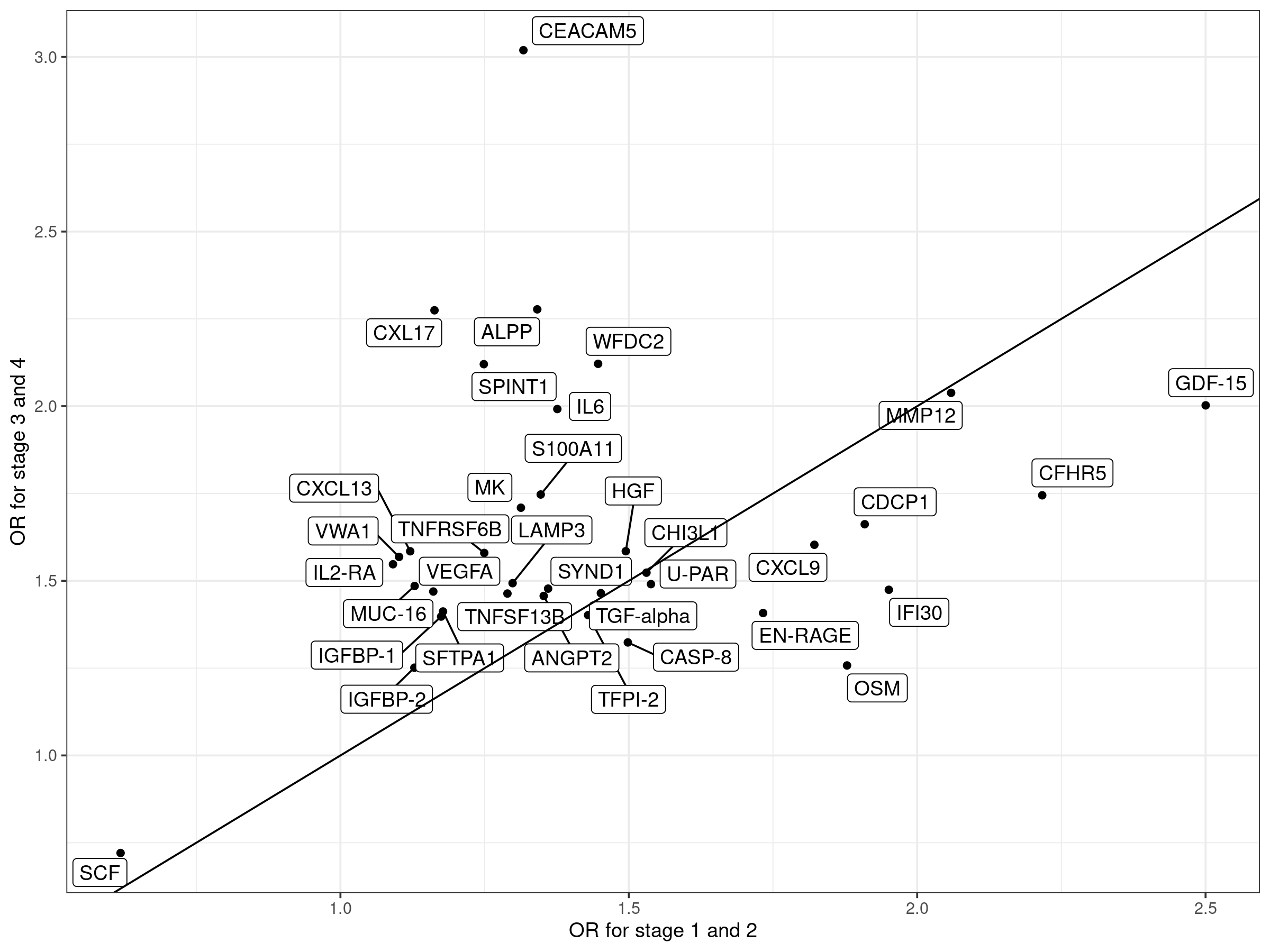

Supplementary Figure 7**:** Heterogeneity in risk-associations for VWA1 and IGFBP-1 by smoking status (current vs former, *p*_het_ <0.05).

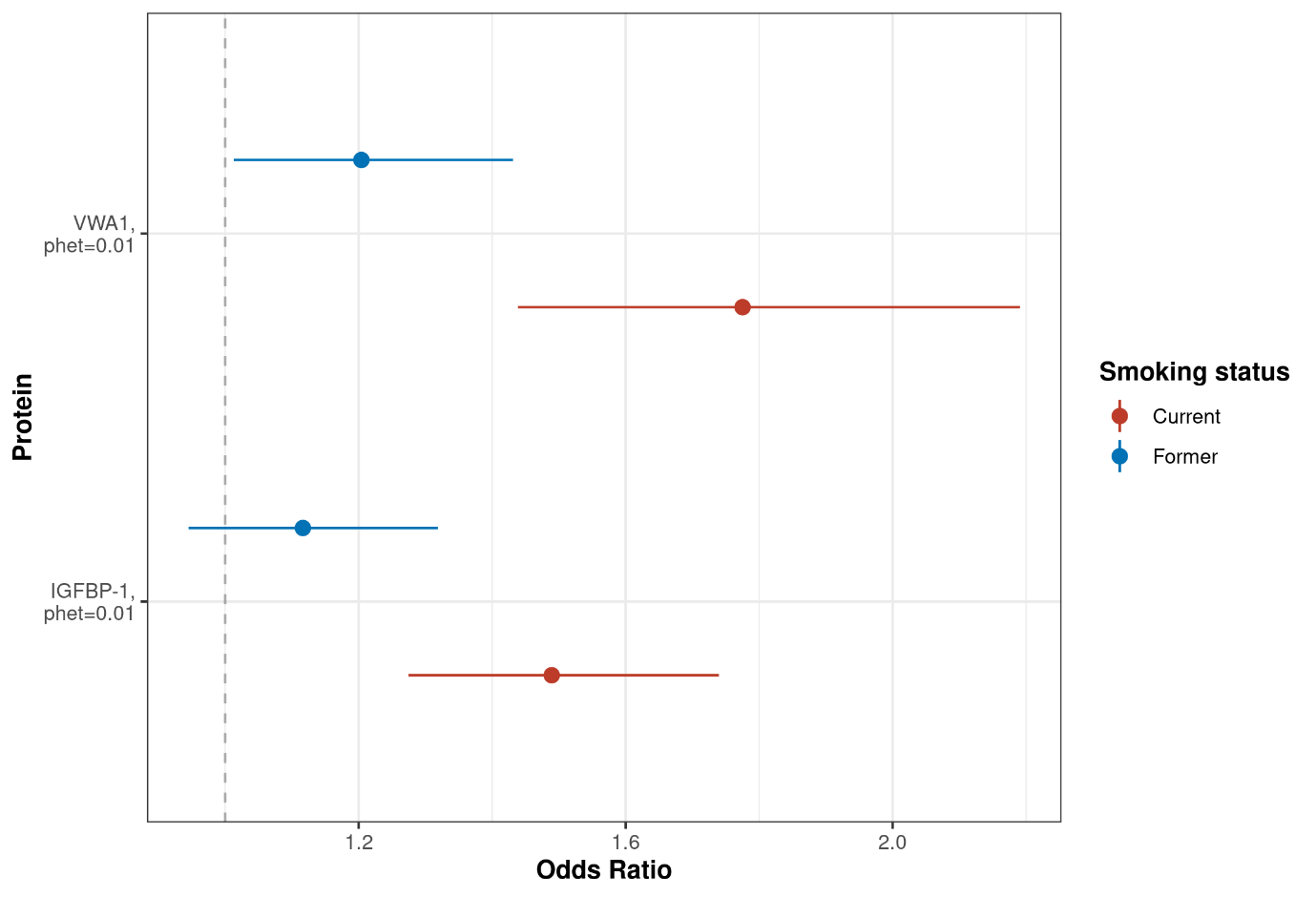

Supplementary Figure 8**:** Heterogeneity in risk-associations for 11 proteins by lead time between blood draw and lung cancer diagnosis (*p*_trend_<0.05).

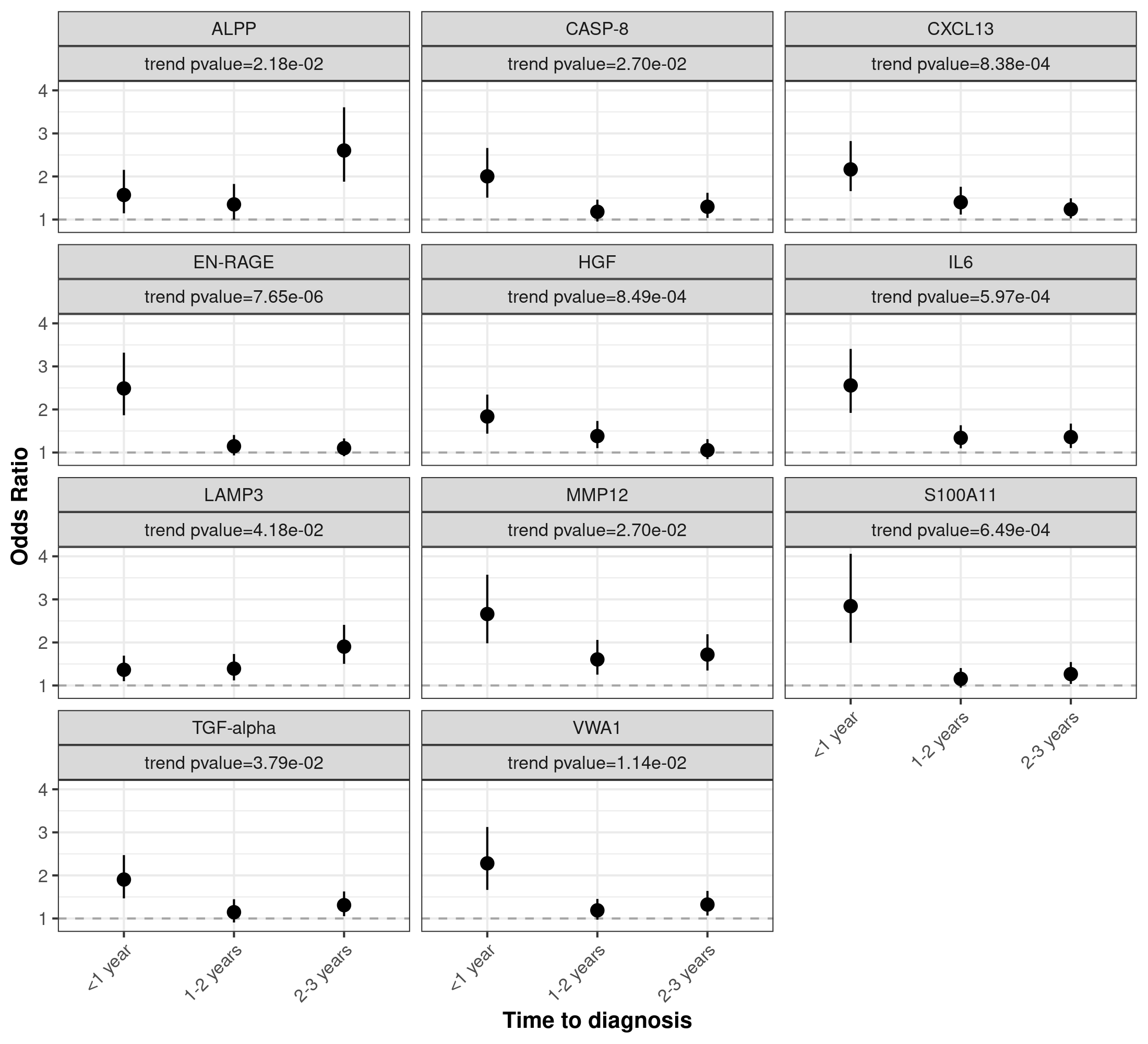

Supplementary Figure 9**:** Heterogeneity in risk-associations for S100A4 and RFNG between the Singapore Chinese Health Study (SCHS) vs. other cohorts from the USA, Europe, and Australia (*p*_het_<0.05).

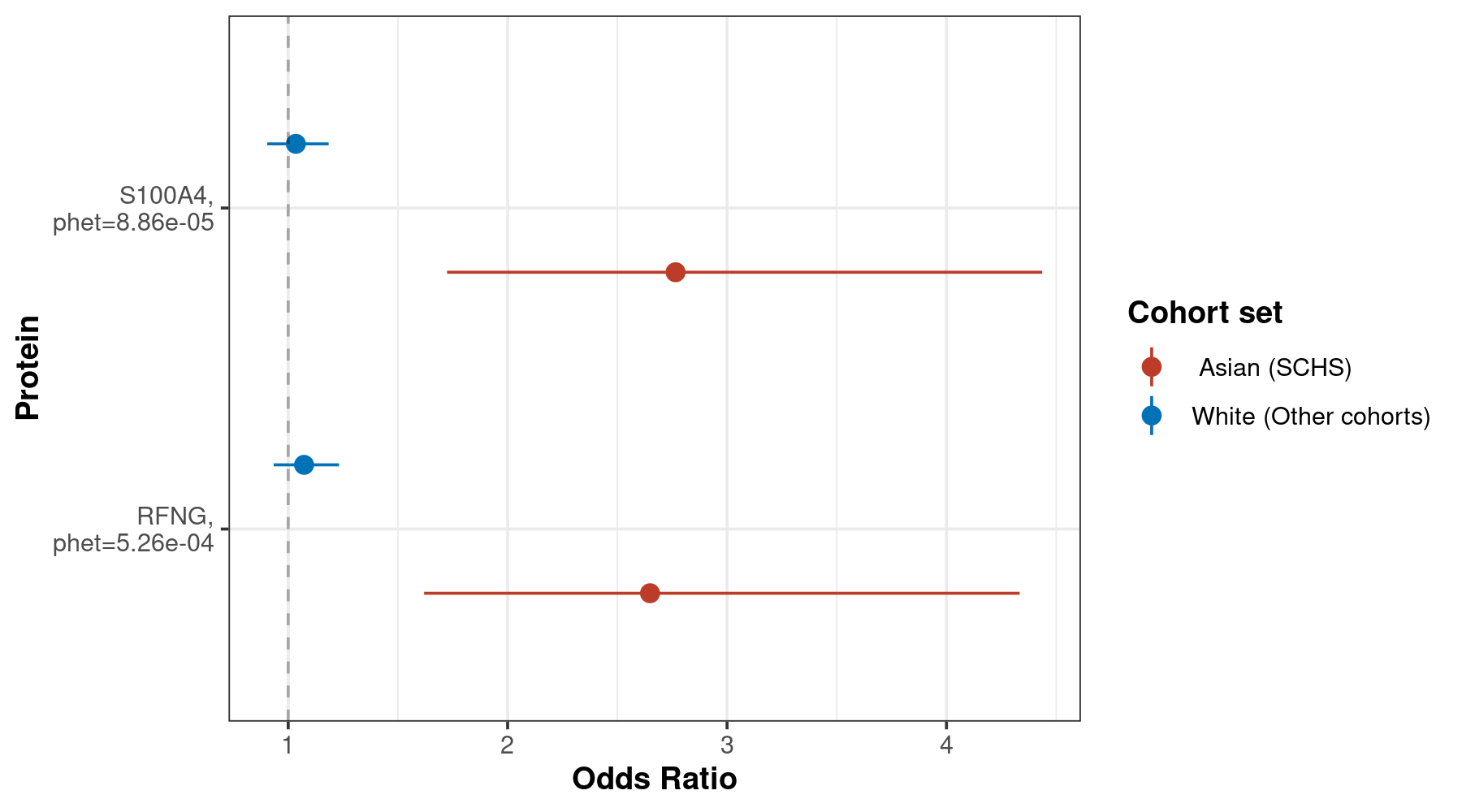

Supplementary Figure 10: Cross-sectional relationships between protein measurements and smoking exposure information among control participants.

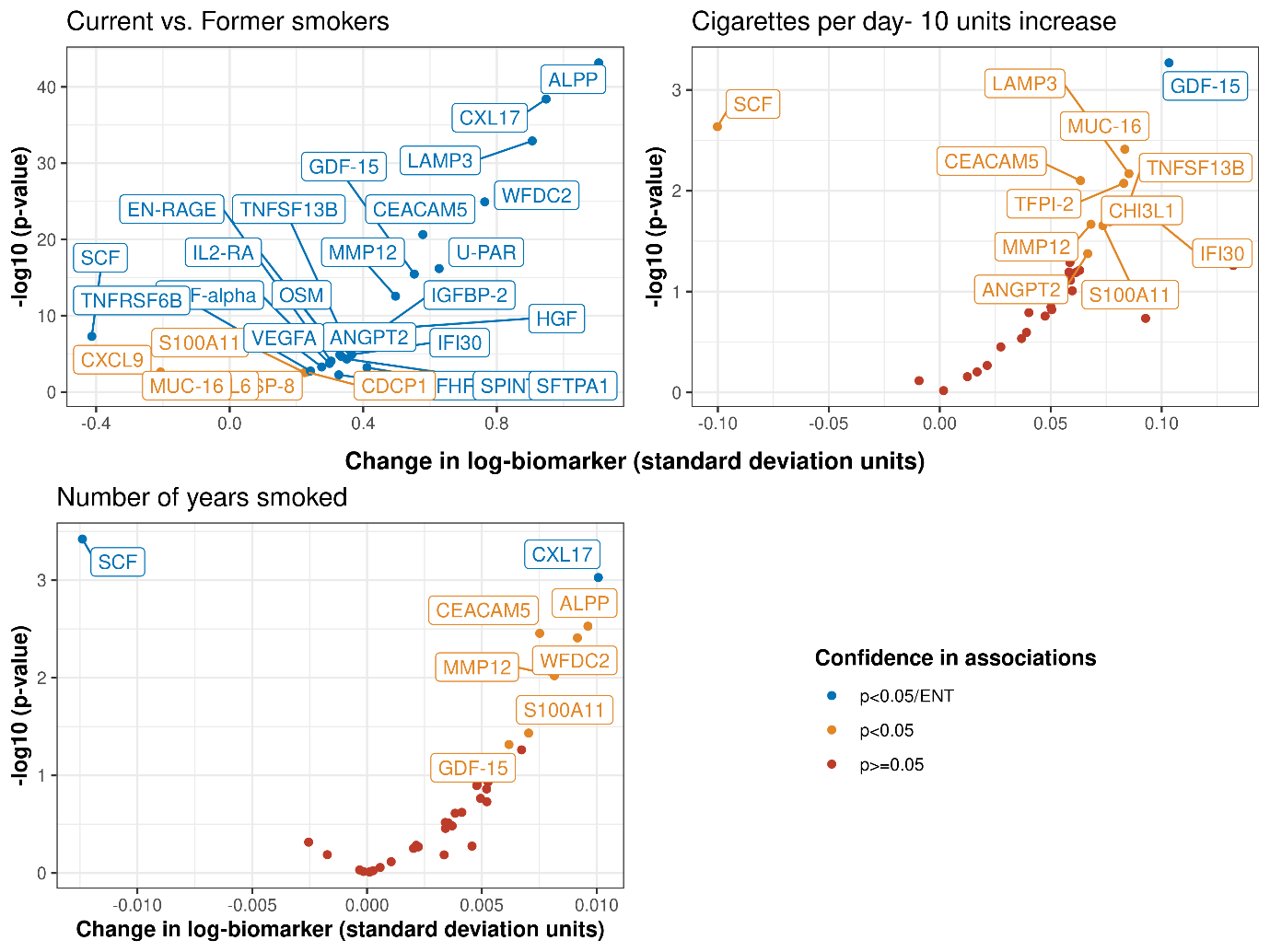

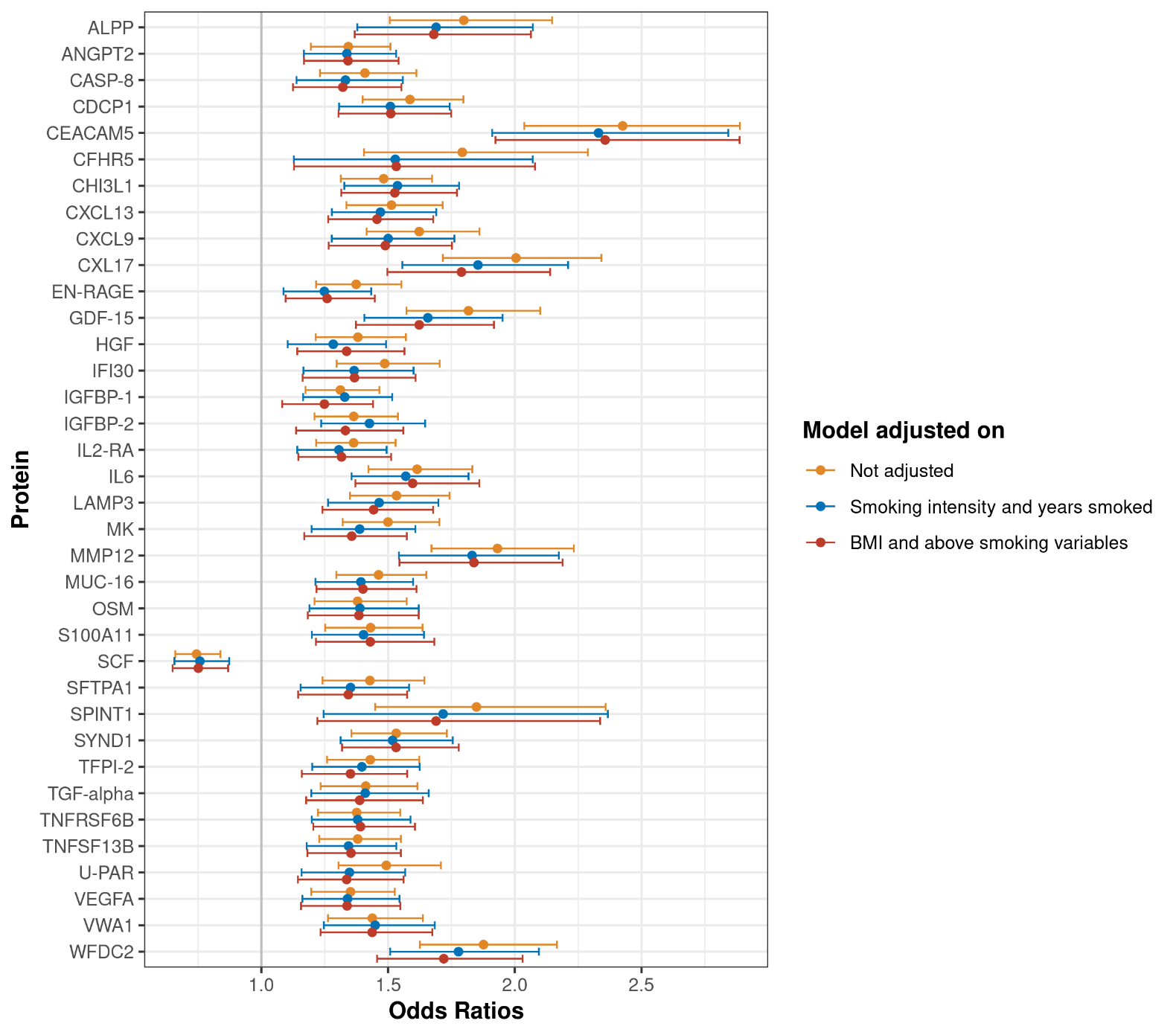
Supplementary Figure 11: Lung cancer odds ratios for the 36 proteins associated with imminent lung cancer before and after detailed adjustment for smoking intensity, duration and BMI.

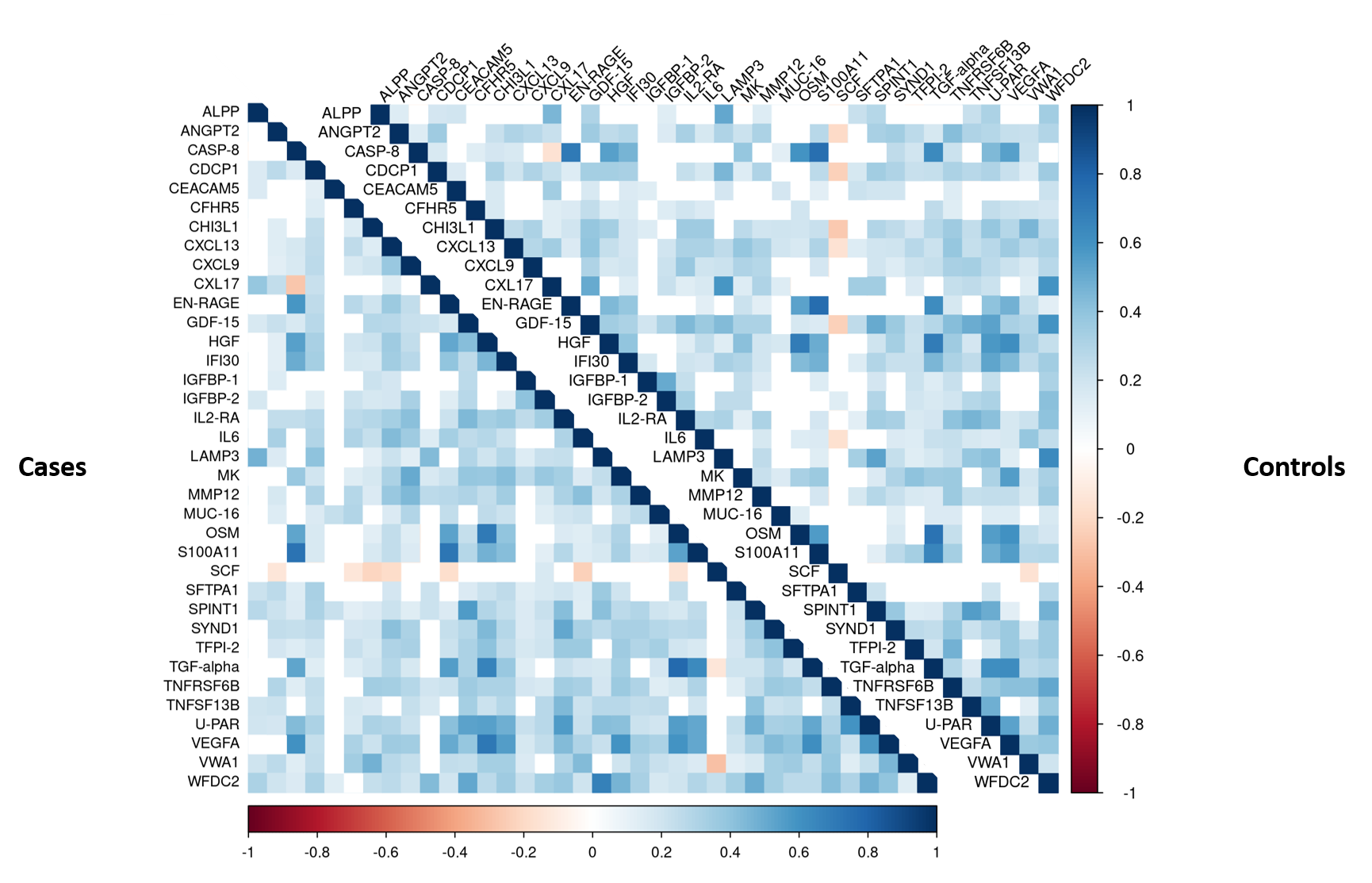
Supplementary Figure 12: Correlation analyses among 36 proteins associated with imminent lung cancer identified among 731 cases and 731 matched controls in the Lung Cancer Cohort Consortium. The figure depicts Pearson’s correlation coefficients between markers separately in cases and controls.

Pearson’s correlation coefficients between markers were estimated after accounting for variation due to sex, age, and cohort (see Supplementary Methods).

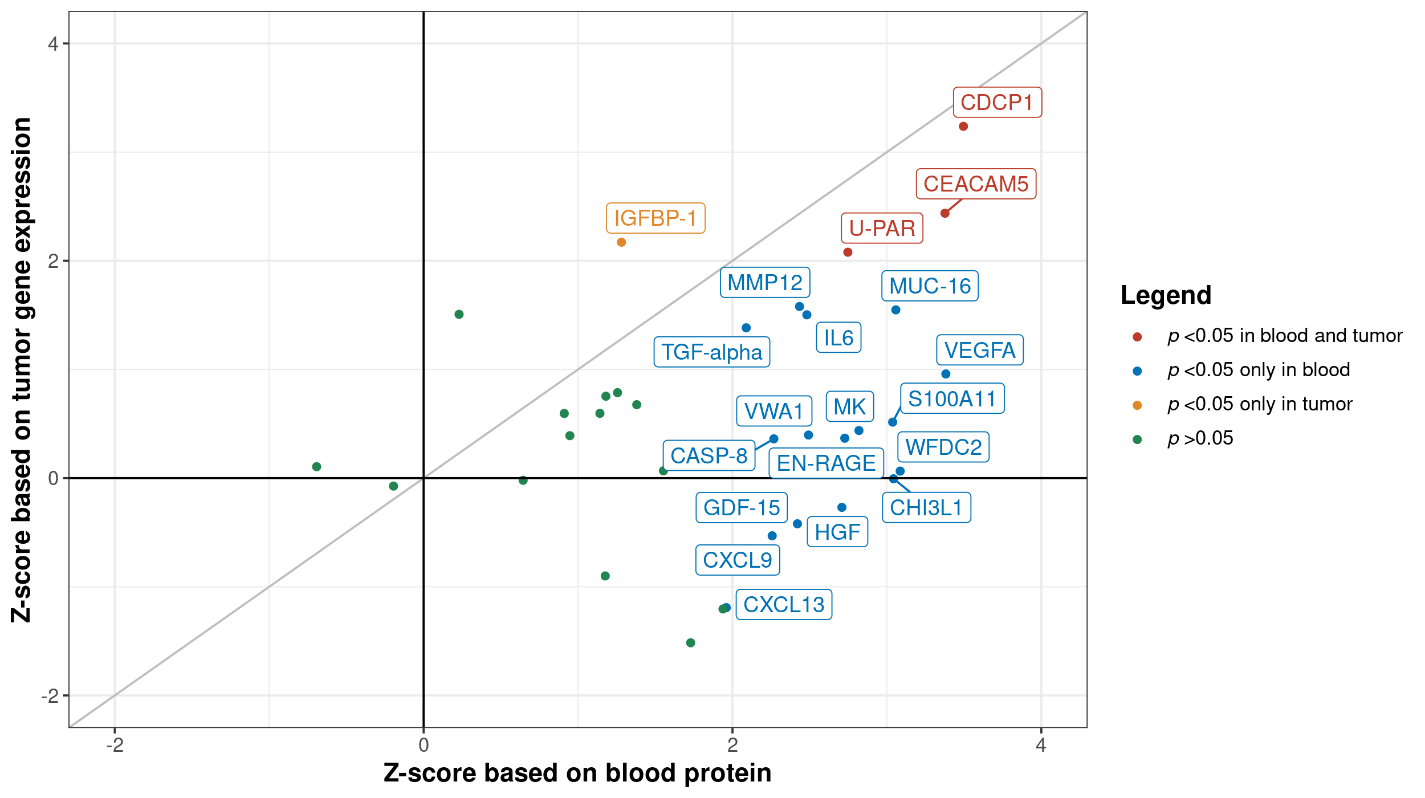
Supplementary Figure 13**:** Association between the 36 markers of imminent lung cancer diagnosis and overall mortality among lung cancer cases, based on direct measurements among participants in the Lung Cancer Cohort Consortium (z-scores on x-axis) and tumor gene expression in TCGA (z-scores on y-axis).

| Among lung cancer cases | N = 731 |
| --- | --- |
| Mortality status |  |
| Alive at end of follow-up | 56 (7.7%) |
| Death | 673 (92%) |
| Lung cancer diagnosis at death* | 2 (0.3%) |

The grey diagonal line on the figure represents identical Z-scores of the association of the biomarkers with all-cause mortality among lung cancer cases when measured in the blood (x-axis) and when their gene expression is measured in the tumour (y-axis).
